# The causal effect of trade unions on workers’ health: A parametric g-formula approach using longitudinal data from the Panel Study of Income Dynamics (PSID)

**DOI:** 10.1101/2025.11.13.25340154

**Authors:** Juan Gonzalez-Hijon, Natasia Hamarat, Theocharis Kromydas, Jacques Wels

## Abstract

**Background:** Trade unions are increasingly recognized as public health actors. They both protect workers’ health by ensuring workplace health and safety and, indirectly, by providing economic advantages to their members. However, previous research has not addressed the direct and indirect causal pathways between trade unions and health and only focused on membership, omitting the important role that the presence of trade union has within the workplace.

**Methods:** We used two decades of nationally representative longitudinal data from the United States (PSID 2001-2021; ≈26,000 individuals; ≈117,000 person-years), used Exploratory Causal Discovery to detect causal pathways and applied parametric g-formula modelling to account for time-varying confounding.

**Findings:** Sustained union membership was associated with lower psychological distress (Mean Ratio – MR 0.944; 95% CI 0.916–0.974), with a persistent direct effect after adjusting for income, working hours, and housing (MR 0.937; 95% CI 0.907–0.968). Always workplace union presence showed an association (MR 0.972; 95% CI 0.945–0.999) that attenuated after adjusting for mediators. Improvements in self-reported health were primarily mediated by income and working-hours pathways (always union membership total adjust MR 0.991; 95% CI 0.983–1.000; always union presence total adjust: MR 0.990; 95% CI 0.982– 0.998).

**Interpretation:** By examining both union membership and workplace union presence, this study captures the individual and institutional dimensions of unionization that have not been simultaneously addressed in prior research. It highlights the significant role trade unions play in improving workers’ health and the threat the erosion of collective bargaining in the US might pose for population health.

## Introduction

### Trade unions are increasingly recognized not only as economic actors but as institutions that protect workers health ^1–5^

An emerging body of literature has documented this relationship, often from two perspectives. First, there is a focus on unions’ role in enforcing workplace safety ^6–9^. Unions are seen as representative institutions that advocate for higher wages ^10^ as well as adequate working conditions and reduced workplace hazards ^11–13^. Second, a more recent perspective considers trade unions as public health actors ^14^, aiming to improve both the physical and mental health of the workforce. This second approach, often relying on longitudinal data, has predominantly focused on trade union membership ^3,15^ – i.e., whether a worker is affiliated with a trade union or representative organization – to address workers’ health change over time. However, it has overlooked the crucial role unions might play in protecting non-unionized workers. Whilst it has long been evidenced that trade unions contribute to raising wages for the unionized workforce ^16–18^ but also for the non-unionized working in unionized workplaces ^19–21^, the health benefits of working in a unionized workplace independently of being unionized have not been properly addressed.

The lack of attention to unions in public health may stem from declining unionization rates. In the US, union density – i.e., trade union membership rates among the working population – have dropped from 35 percent in the early 1960s to around 12 percent in the 2020s ^22^. Changes in workers’ attitudes towards unions explain little of this trend and a large part is due to change in industry ^23^. One major issue is that, in the US, any worker who wish to unionize must first secure the support of 50 percent of their coworkers, rather than being able to join a union individually ^22^. At the company level, union presence depends on whether workers have chosen to unionize through a formal election overseen by the National Labor Relations Board (NLRB) or through voluntary recognition by the employer. Once a union is established in a workplace, it becomes the exclusive bargaining representative for all employees in the bargaining unit. In other words, the possibility to unionize is a key determinant of union membership. Studies have found that declining organizing efforts and success account for a large part of the decline in union membership ^23,24^ indicating the necessity to move beyond a membership approach.

Examining the relationship between trade unions and workers’ health requires establishing causation – both in terms of the directionality of the observed relationship and the pathways through which unions influence health.

When assessing whether unions affect workers’ health, one should account for reverse causation as union membership is often used as a voice in response to poor job conditions ^16,26,27^. But the decision to unionize is not only affected by working conditions. For instance, it was evidenced that changes in work characteristics from manual versus non-manual work have plaid a role in explaining declining trends ^28^. These factors come together with changes in legislations and political administrations that have influenced the recognition of workplace unions ^29^. In other words, union membership is affected by multiple parameters whilst union presence, although not at random, is affected by social change rather than workers’ voices. Another issue that over-controlling for certain variables might bias causal estimates. Previous studies on the relationship between trade unions and workers’ health were adjusted for wages ^1,3^. Whilst this allows to calculate the total effect, it somehow blocks the pathway between trade unions and health because labor incomes are part of the explanatory mechanism ^30^ as the presence of workplace collective bargaining – not union membership as such ^31^ – is a driver of increased wages^31^ and wages have effects on health on physical and mental health^32^. Although, things have slightly changed over the recent period ^33^ and differences exist when looking at genders ^34^, wages (as well as financial variables derived from wages) play a mediating role: unions affect wages and health, and wages affect health outcomes. This highlights the need to not over adjust models and distinguish between total (without incomes controls) and direct effects (with incomes controls).

Using longitudinal US data, this study addresses three research questions:

RQ1: Is there a causal relationship between the presence of trade union within the workplace and mental health (Kesseler-6 psychological distress score) / self-reported health outcomes?
RQ2: Does the nature of this relationship varies depending on the levels of adjustment of the model?
RQ3: Is there a difference in mental health and self-reported physical health between employees who are union members and employees who work in a workplace with union presence (regardless of individual membership)?

## Methods

### Registered protocol

The study protocol was pre-registered on the Open Science Framework (OSF; https://doi.org/10.17605/OSF.IO/QDXTF).

### Data Source and Study Population

We used data from the PSID Individual and Family files. The PSID (Panel Study of Income Dynamics), an U.S. longitudinal household survey, collected data via annual interviews from 1968 to 1997 and biennially thereafter. We constructed a panel dataset with stable cross-wave identifiers and used person-year observations as analysis unit. The analytic sample included all person-year observations with complete outcome data; responders aged <18 were excluded. To ensure comparability between outcomes, analyses were restricted to the overlapping period 2001–2021. The total sample includes ≈26 000 individuals (≈117 000 persons-years).

### Measures

#### Independent Variables

Our primary independent variables were: 1) Presence of union at work, a binary indicator (1=present, 0=not present); and 2) Union membership, a binary indicator (1=member, 0=not a member).

#### Outcome Variables

1) Self-reported health was measured with the question, “Would you say your health in general is excellent, very good, good, fair, or poor?” scored from 1 (excellent) to 5 (poor). 2) Mental health was measured using the Kessler-6 (K-6) scale, a validated screener for nonspecific psychological distress over the past 30 days (Cronbach’s alphaL=L0.89), with scores ranging from 0 to 24 (higher scores indicate greater distress)^35,36^.

#### Covariates

Age (in years), Gender, Educational Level, Region of residence (grouped into six geographic regions), Industry, Working Hours (log-transformed and cantered), Housing tenure, and Family income (OECD-equivalized household income, log-transformed and centered)^37^, calendar Year and survey Wave; reference categories are shown on Table 1. Employment Status was treated deterministically to exclude non-employed waves from parametric g-formula simulation steps but retaining observations to preserve temporal continuity of the panel.

**Table 1.** Descriptive statistics. The category labelled “[Ref.]” was used as the reference group in the analyses.

|  | Baseline<br>Union Presence at Workplace |  | Baseline<br>Union Membership |  |
| --- | --- | --- | --- | --- |
|  | Not Union<br>Presence<br>(N = 21771) | Union<br>Presence<br>(N = 4464) | Not Union<br>Membership<br>(N = 22486) | Union<br>Membership<br>(N = 3749) |
| **Gender** |  |  |  |  |
| Female | 5506 (25.3%) | 1033 (23.1%) | 5743 (25.5%) | 796 (21.2%) |
| Male [Ref.] | 16265 (74.7%) | 3431 (76.9%) | 16743 (74.5%) | 2953 (78.8%) |
| **Age at Baseline** |  |  |  |  |
| 18-24 | 8395 (38.6%) | 1691 (37.9%) | 8675 (38.6%) | 1411 (37.6%) |
| 25-34 | 5368 (24.7%) | 898 (20.1%) | 5545 (24.7%) | 721 (19.2%) |
| 35-44 | 3731 (17.1%) | 853 (19.1%) | 3858 (17.2%) | 726 (19.4%) |
| 45-54 | 2567 (11.8%) | 689 (15.4%) | 2654 (11.8%) | 602 (16.1%) |
| 55-64 | 1114 (5.1%) | 212 (4.7%) | 1145 (5.1%) | 181 (4.8%) |
| 65+ | 596 (2.7%) | 121 (2.7%) | 609 (2.7%) | 108 (2.9%) |
| **Race** |  |  |  |  |
| White | 12228 (56.2%) | 2175 (48.7%) | 12479 (55.5%) | 1924 (51.3%) |
| Other [Ref.] | 9220 (42.4%) | 2253 (50.5%) | 9676 (43.0%) | 1797 (47.9%) |
| Missing | 323 (1.5%) | 36 (0.8%) | 331 (1.5%) | 28 (0.7%) |
| **Self-Reported Health** |  |  |  |  |
| Mean (SD) | 2.30 (0.982) | 2.32 (0.969) | 2.30 (0.981) | 2.32 (0.969) |
| Median [Min, Max] | 2.00 [1.00, 5.00] | 2.00 [1.00, 5.00] | 2.00 [1.00, 5.00] | 2.00 [1.00, 5.00] |
| **Mental Health** |  |  |  |  |
| Mean (SD) | 3.47 (3.90) | 3.16 (3.65) | 3.48 (3.91) | 3.04 (3.52) |
| Median [Min, Max] | 2.00 [0, 24.0] | 2.00 [0, 24.0] | 2.00 [0, 24.0] | 2.00 [0, 24.0] |
| Missing | 1771 (8.1%) | 292 (6.5%) | 1817 (8.1%) | 246 (6.6%) |
| **Employment Status** |  |  |  |  |
| Employed | 20148 (92.5%) | 4464 (100%) | 20863 (92.8%) | 3749 (100%) |
| Not Employed | 1623 (7.5%) | 0 (0%) | 1623 (7.2%) | 0 (0%) |
| **Equivalized Family Income** |  |  |  |  |
| Mean (SD) | 41200 (54500) | 42300 (29600) | 40900 (53800) | 44100 (30300) |
| Median [Min, Max] | 30000 [0, 2630000] | 36500 [0, 543000] | 30000 [0, 2630000] | 38000 [0, 543000] |
| **Family Members in Household** |  |  |  |  |
| Mean (SD) | 3.26 (1.59) | 3.41 (1.57) | 3.26 (1.59) | 3.45 (1.58) |
| Median [Min, Max] | 3.00 [1.00, 16.0] | 3.00 [1.00, 13.0] | 3.00 [1.00, 16.0] | 3.00 [1.00, 13.0] |
| **Working Hours** |  |  |  |  |
| Mean (SD) | 42.9 (11.5) | 42.8 (10.1) | 42.9 (11.5) | 43.1 (9.87) |
| Median [Min, Max] | 40.0 [1.00, 97.0] | 40.0 [1.00, 98.0] | 40.0 [1.00, 97.0] | 40.0 [6.00, 98.0] |
| Missing | 469 (2.2%) | 52 (1.2%) | 495 (2.2%) | 26 (0.7%) |
| **Housing Tenure** |  |  |  |  |
| Owns House | 11235 (51.6%) | 2719 (60.9%) | 11561 (51.4%) | 2393 (63.8%) |
| Pays Rent or Other [Ref.] | 10536 (48.4%) | 1745 (39.1%) | 10925 (48.6%) | 1356 (36.2%) |
| **Education** |  |  |  |  |
| Elementary School [Ref.] | 141 (0.6%) | 36 (0.8%) | 145 (0.6%) | 32 (0.9%) |
| Middle School | 10190 (46.8%) | 2216 (49.6%) | 10550 (46.9%) | 1856 (49.5%) |
| High School | 366 (1.7%) | 65 (1.5%) | 378 (1.7%) | 53 (1.4%) |
| University (No-Degree) | 4598 (21.1%) | 1013 (22.7%) | 4757 (21.2%) | 854 (22.8%) |
| University (Degree) | 3360 (15.4%) | 602 (13.5%) | 3464 (15.4%) | 498 (13.3%) |
| Non-US System | 1878 (8.6%) | 330 (7.4%) | 1923 (8.6%) | 285 (7.6%) |
| Missing | 1238 (5.7%) | 202 (4.5%) | 1269 (5.6%) | 171 (4.6%) |
| **Industry** |  |  |  |  |
| Accommodation and Food Services | 1319 (6.1%) | 96 (2.2%) | 1347 (6.0%) | 68 (1.8%) |
| Agriculture, Forestry, Fisheries and Mining | 672 (3.1%) | 67 (1.5%) | 690 (3.1%) | 49 (1.3%) |
| Business and Repair Services | 2207 (10.1%) | 129 (2.9%) | 2249 (10.0%) | 87 (2.3%) |
| Construction | 1864 (8.6%) | 379 (8.5%) | 1912 (8.5%) | 331 (8.8%) |
| Entertainment and Recreation Services | 345 (1.6%) | 32 (0.7%) | 351 (1.6%) | 26 (0.7%) |
| Finance, Insurance, and Real Estate | 1296 (6.0%) | 69 (1.5%) | 1313 (5.8%) | 52 (1.4%) |
| Health Care and Social Assistance | 1562 (7.2%) | 248 (5.6%) | 1629 (7.2%) | 181 (4.8%) |
| Information Services | 288 (1.3%) | 55 (1.2%) | 293 (1.3%) | 50 (1.3%) |
| Manufacturing [Ref.] | 3691 (17.0%) | 854 (19.1%) | 3808 (16.9%) | 737 (19.7%) |
| Personal Services | 295 (1.4%) | 146 (3.3%) | 299 (1.3%) | 142 (3.8%) |
| Professional, Scientific, Technical and Educational Services | 1811 (8.3%) | 797 (17.9%) | 1928 (8.6%) | 680 (18.1%) |
| Public Administration | 933 (4.3%) | 533 (11.9%) | 1042 (4.6%) | 424 (11.3%) |
| Transportation, Communications, and other Public Utilities | 1526 (7.0%) | 734 (16.4%) | 1606 (7.1%) | 654 (17.4%) |
| Wholesale and Retail Trade | 3851 (17.7%) | 315 (7.1%) | 3904 (17.4%) | 262 (7.0%) |
| Missing | 111 (0.5%) | 10 (0.2%) | 115 (0.5%) | 6 (0.2%) |
| **Region of residence** |  |  |  |  |
| Midsouth Atlantic | 7287 (33.5%) | 1584 (35.5%) | 7587 (33.7%) | 1284 (34.2%) |
| Midwest | 5238 (24.1%) | 1241 (27.8%) | 5392 (24.0%) | 1087 (29.0%) |
| New England [Ref.] | 652 (3.0%) | 142 (3.2%) | 658 (2.9%) | 136 (3.6%) |
| Pacific | 2795 (12.8%) | 744 (16.7%) | 2870 (12.8%) | 669 (17.8%) |
| South | 4403 (20.2%) | 548 (12.3%) | 4540 (20.2%) | 411 (11.0%) |
| West | 1317 (6.0%) | 176 (3.9%) | 1348 (6.0%) | 145 (3.9%) |
| Other | 79 (0.4%) | 29 (0.6%) | 91 (0.4%) | 17 (0.5%) |
| **Recruitment Year** |  |  |  |  |
| 2001 - 2004 | 9659 (44.4%) | 2398 (53.7%) | 9969 (44.3%) | 2088 (55.7%) |
| 2005 - 2009 | 4257 (19.6%) | 776 (17.4%) | 4359 (19.4%) | 674 (18.0%) |
| 2010 - 2014 | 2550 (11.7%) | 404 (9.1%) | 2638 (11.7%) | 316 (8.4%) |
| 2015 - 2021 | 5305 (24.4%) | 886 (19.8%) | 5520 (24.5%) | 671 (17.9%) |
|  | Not Union<br>Presence<br>(N = 21771) | Union<br>Presence<br>(N = 4464) | Not Union<br>Membership<br>(N = 22486) | Union<br>Membership<br>(N = 3749) |
| **Gender** |  |  |  |  |
| Female | 5506 (25.3%) | 1033 (23.1%) | 5743 (25.5%) | 796 (21.2%) |
| Male [Ref.] | 16265 (74.7%) | 3431 (76.9%) | 16743 (74.5%) | 2953 (78.8%) |
| **Age at Baseline** |  |  |  |  |
| 18-24 | 8395 (38.6%) | 1691 (37.9%) | 8675 (38.6%) | 1411 (37.6%) |
| 25-34 | 5368 (24.7%) | 898 (20.1%) | 5545 (24.7%) | 721 (19.2%) |
| 35-44 | 3731 (17.1%) | 853 (19.1%) | 3858 (17.2%) | 726 (19.4%) |
| 45-54 | 2567 (11.8%) | 689 (15.4%) | 2654 (11.8%) | 602 (16.1%) |
| 55-64 | 1114 (5.1%) | 212 (4.7%) | 1145 (5.1%) | 181 (4.8%) |
| 65+ | 596 (2.7%) | 121 (2.7%) | 609 (2.7%) | 108 (2.9%) |
| **Race** |  |  |  |  |
| White | 12228 (56.2%) | 2175 (48.7%) | 12479 (55.5%) | 1924 (51.3%) |
| Other [Ref.] | 9220 (42.4%) | 2253 (50.5%) | 9676 (43.0%) | 1797 (47.9%) |
| Missing | 323 (1.5%) | 36 (0.8%) | 331 (1.5%) | 28 (0.7%) |
| **Self-Reported Health** |  |  |  |  |
| Mean (SD) | 2.30 (0.982) | 2.32 (0.969) | 2.30 (0.981) | 2.32 (0.969) |
| Median [Min, Max] | 2.00 [1.00, 5.00] | 2.00 [1.00, 5.00] | 2.00 [1.00, 5.00] | 2.00 [1.00, 5.00] |
| **Mental Health** |  |  |  |  |
| Mean (SD) | 3.47 (3.90) | 3.16 (3.65) | 3.48 (3.91) | 3.04 (3.52) |
| Median [Min, Max] | 2.00 [0, 24.0] | 2.00 [0, 24.0] | 2.00 [0, 24.0] | 2.00 [0, 24.0] |
| Missing | 1771 (8.1%) | 292 (6.5%) | 1817 (8.1%) | 246 (6.6%) |
| **Employment Status** |  |  |  |  |
| Employed | 20148 (92.5%) | 4464 (100%) | 20863 (92.8%) | 3749 (100%) |
| Not Employed | 1623 (7.5%) | 0 (0%) | 1623 (7.2%) | 0 (0%) |
| **Equivalized Family Income** |  |  |  |  |
| Mean (SD) | 41200 (54500) | 42300 (29600) | 40900 (53800) | 44100 (30300) |
| Median [Min, Max] | 30000 [0, 2630000] | 36500 [0, 543000] | 30000 [0, 2630000] | 38000 [0, 543000] |
| **Family Members in Household** |  |  |  |  |
| Mean (SD) | 3.26 (1.59) | 3.41 (1.57) | 3.26 (1.59) | 3.45 (1.58) |
| Median [Min, Max] | 3.00 [1.00, 16.0] | 3.00 [1.00, 13.0] | 3.00 [1.00, 16.0] | 3.00 [1.00, 13.0] |
| **Working Hours** |  |  |  |  |
| Mean (SD) | 42.9 (11.5) | 42.8 (10.1) | 42.9 (11.5) | 43.1 (9.87) |
| Median [Min, Max] | 40.0 [1.00, 97.0] | 40.0 [1.00, 98.0] | 40.0 [1.00, 97.0] | 40.0 [6.00, 98.0] |
| Missing | 469 (2.2%) | 52 (1.2%) | 495 (2.2%) | 26 (0.7%) |
| **Housing Tenure** |  |  |  |  |
| Owns House | 11235 (51.6%) | 2719 (60.9%) | 11561 (51.4%) | 2393 (63.8%) |
| Pays Rent or Other [Ref.] | 10536 (48.4%) | 1745 (39.1%) | 10925 (48.6%) | 1356 (36.2%) |
| **Education** |  |  |  |  |
| Elementary School [Ref.] | 141 (0.6%) | 36 (0.8%) | 145 (0.6%) | 32 (0.9%) |
| Middle School | 10190 (46.8%) | 2216 (49.6%) | 10550 (46.9%) | 1856 (49.5%) |
| High School | 366 (1.7%) | 65 (1.5%) | 378 (1.7%) | 53 (1.4%) |
| University (No-Degree) | 4598 (21.1%) | 1013 (22.7%) | 4757 (21.2%) | 854 (22.8%) |
| University (Degree) | 3360 (15.4%) | 602 (13.5%) | 3464 (15.4%) | 498 (13.3%) |
| Non-US System | 1878 (8.6%) | 330 (7.4%) | 1923 (8.6%) | 285 (7.6%) |
| Missing | 1238 (5.7%) | 202 (4.5%) | 1269 (5.6%) | 171 (4.6%) |
| **Industry** |  |  |  |  |
| Accommodation and Food Services | 1319 (6.1%) | 96 (2.2%) | 1347 (6.0%) | 68 (1.8%) |
| Agriculture, Forestry, Fisheries and Mining | 672 (3.1%) | 67 (1.5%) | 690 (3.1%) | 49 (1.3%) |
| Business and Repair Services | 2207 (10.1%) | 129 (2.9%) | 2249 (10.0%) | 87 (2.3%) |
| Construction | 1864 (8.6%) | 379 (8.5%) | 1912 (8.5%) | 331 (8.8%) |
| Entertainment and Recreation Services | 345 (1.6%) | 32 (0.7%) | 351 (1.6%) | 26 (0.7%) |
| Finance, Insurance, and Real Estate | 1296 (6.0%) | 69 (1.5%) | 1313 (5.8%) | 52 (1.4%) |
| Health Care and Social Assistance | 1562 (7.2%) | 248 (5.6%) | 1629 (7.2%) | 181 (4.8%) |
| Information Services | 288 (1.3%) | 55 (1.2%) | 293 (1.3%) | 50 (1.3%) |
| Manufacturing [Ref.] | 3691 (17.0%) | 854 (19.1%) | 3808 (16.9%) | 737 (19.7%) |
| Personal Services | 295 (1.4%) | 146 (3.3%) | 299 (1.3%) | 142 (3.8%) |
| Professional, Scientific, Technical and Educational Services | 1811 (8.3%) | 797 (17.9%) | 1928 (8.6%) | 680 (18.1%) |
| Public Administration | 933 (4.3%) | 533 (11.9%) | 1042 (4.6%) | 424 (11.3%) |
| Transportation, Communications, and other Public Utilities | 1526 (7.0%) | 734 (16.4%) | 1606 (7.1%) | 654 (17.4%) |
| Wholesale and Retail Trade | 3851 (17.7%) | 315 (7.1%) | 3904 (17.4%) | 262 (7.0%) |
| Missing | 111 (0.5%) | 10 (0.2%) | 115 (0.5%) | 6 (0.2%) |
| **Region of residence** |  |  |  |  |
| Midsouth Atlantic | 7287 (33.5%) | 1584 (35.5%) | 7587 (33.7%) | 1284 (34.2%) |
| Midwest | 5238 (24.1%) | 1241 (27.8%) | 5392 (24.0%) | 1087 (29.0%) |
| New England [Ref.] | 652 (3.0%) | 142 (3.2%) | 658 (2.9%) | 136 (3.6%) |
| Pacific | 2795 (12.8%) | 744 (16.7%) | 2870 (12.8%) | 669 (17.8%) |
| South | 4403 (20.2%) | 548 (12.3%) | 4540 (20.2%) | 411 (11.0%) |
| West | 1317 (6.0%) | 176 (3.9%) | 1348 (6.0%) | 145 (3.9%) |
| Other | 79 (0.4%) | 29 (0.6%) | 91 (0.4%) | 17 (0.5%) |
| **Recruitment Year** |  |  |  |  |
| 2001 - 2004 | 9659 (44.4%) | 2398 (53.7%) | 9969 (44.3%) | 2088 (55.7%) |
| 2005 - 2009 | 4257 (19.6%) | 776 (17.4%) | 4359 (19.4%) | 674 (18.0%) |
| 2010 - 2014 | 2550 (11.7%) | 404 (9.1%) | 2638 (11.7%) | 316 (8.4%) |

| 2015 - 2021 | 5305 (24.4%) | 886 (19.8%) | 5520 (24.5%) | 671 (17.9%) |
| --- | --- | --- | --- | --- |
|  | Baseline<br>Union Presence at Workplace |  | Baseline<br>Union Membership |  |
|  | Not Union<br>Presence<br>(N = 21771) | Union<br>Presence<br>(N = 4464) | Not Union<br>Membership<br>(N = 22486) | Union<br>Membership<br>(N = 3749) |
| **Gender** |  |  |  |  |
| Female | 5506 (25.3%) | 1033 (23.1%) | 5743 (25.5%) | 796 (21.2%) |
| Male [Ref.] | 16265 (74.7%) | 3431 (76.9%) | 16743 (74.5%) | 2953 (78.8%) |
| **Age at Baseline** |  |  |  |  |
| 18-24 | 8395 (38.6%) | 1691 (37.9%) | 8675 (38.6%) | 1411 (37.6%) |
| 25-34 | 5368 (24.7%) | 898 (20.1%) | 5545 (24.7%) | 721 (19.2%) |
| 35-44 | 3731 (17.1%) | 853 (19.1%) | 3858 (17.2%) | 726 (19.4%) |
| 45-54 | 2567 (11.8%) | 689 (15.4%) | 2654 (11.8%) | 602 (16.1%) |
| 55-64 | 1114 (5.1%) | 212 (4.7%) | 1145 (5.1%) | 181 (4.8%) |
| 65+ | 596 (2.7%) | 121 (2.7%) | 609 (2.7%) | 108 (2.9%) |
| **Race** |  |  |  |  |
| White | 12228 (56.2%) | 2175 (48.7%) | 12479 (55.5%) | 1924 (51.3%) |
| Other [Ref.] | 9220 (42.4%) | 2253 (50.5%) | 9676 (43.0%) | 1797 (47.9%) |
| Missing | 323 (1.5%) | 36 (0.8%) | 331 (1.5%) | 28 (0.7%) |
| **Self-Reported Health** |  |  |  |  |
| Mean (SD) | 2.30 (0.982) | 2.32 (0.969) | 2.30 (0.981) | 2.32 (0.969) |
| Median [Min, Max] | 2.00 [1.00, 5.00] | 2.00 [1.00, 5.00] | 2.00 [1.00, 5.00] | 2.00 [1.00, 5.00] |
| **Mental Health** |  |  |  |  |
| Mean (SD) | 3.47 (3.90) | 3.16 (3.65) | 3.48 (3.91) | 3.04 (3.52) |
| Median [Min, Max] | 2.00 [0, 24.0] | 2.00 [0, 24.0] | 2.00 [0, 24.0] | 2.00 [0, 24.0] |
| Missing | 1771 (8.1%) | 292 (6.5%) | 1817 (8.1%) | 246 (6.6%) |
| **Employment Status** |  |  |  |  |
| Employed | 20148 (92.5%) | 4464 (100%) | 20863 (92.8%) | 3749 (100%) |
| Not Employed | 1623 (7.5%) | 0 (0%) | 1623 (7.2%) | 0 (0%) |
| **Equivalized Family Income** |  |  |  |  |
| Mean (SD) | 41200 (54500) | 42300 (29600) | 40900 (53800) | 44100 (30300) |
| Median [Min, Max] | 30000 [0, 2630000] | 36500 [0, 543000] | 30000 [0, 2630000] | 38000 [0, 543000] |
| **Family Members in Household** |  |  |  |  |
| Mean (SD) | 3.26 (1.59) | 3.41 (1.57) | 3.26 (1.59) | 3.45 (1.58) |
| Median [Min, Max] | 3.00 [1.00, 16.0] | 3.00 [1.00, 13.0] | 3.00 [1.00, 16.0] | 3.00 [1.00, 13.0] |
| **Working Hours** |  |  |  |  |
| Mean (SD) | 42.9 (11.5) | 42.8 (10.1) | 42.9 (11.5) | 43.1 (9.87) |
| Median [Min, Max] | 40.0 [1.00, 97.0] | 40.0 [1.00, 98.0] | 40.0 [1.00, 97.0] | 40.0 [6.00, 98.0] |
| Missing | 469 (2.2%) | 52 (1.2%) | 495 (2.2%) | 26 (0.7%) |
| **Housing Tenure** |  |  |  |  |
| Owns House | 11235 (51.6%) | 2719 (60.9%) | 11561 (51.4%) | 2393 (63.8%) |
| Pays Rent or Other [Ref.] | 10536 (48.4%) | 1745 (39.1%) | 10925 (48.6%) | 1356 (36.2%) |
| **Education** |  |  |  |  |
| Elementary School [Ref.] | 141 (0.6%) | 36 (0.8%) | 145 (0.6%) | 32 (0.9%) |
| Middle School | 10190 (46.8%) | 2216 (49.6%) | 10550 (46.9%) | 1856 (49.5%) |
| High School | 366 (1.7%) | 65 (1.5%) | 378 (1.7%) | 53 (1.4%) |
| University (No-Degree) | 4598 (21.1%) | 1013 (22.7%) | 4757 (21.2%) | 854 (22.8%) |
| University (Degree) | 3360 (15.4%) | 602 (13.5%) | 3464 (15.4%) | 498 (13.3%) |
| Non-US System | 1878 (8.6%) | 330 (7.4%) | 1923 (8.6%) | 285 (7.6%) |
| Missing | 1238 (5.7%) | 202 (4.5%) | 1269 (5.6%) | 171 (4.6%) |
| **Industry** |  |  |  |  |
| Accommodation and Food Services | 1319 (6.1%) | 96 (2.2%) | 1347 (6.0%) | 68 (1.8%) |
| Agriculture, Forestry, Fisheries and Mining | 672 (3.1%) | 67 (1.5%) | 690 (3.1%) | 49 (1.3%) |
| Business and Repair Services | 2207 (10.1%) | 129 (2.9%) | 2249 (10.0%) | 87 (2.3%) |
| Construction | 1864 (8.6%) | 379 (8.5%) | 1912 (8.5%) | 331 (8.8%) |
| Entertainment and Recreation Services | 345 (1.6%) | 32 (0.7%) | 351 (1.6%) | 26 (0.7%) |
| Finance, Insurance, and Real Estate | 1296 (6.0%) | 69 (1.5%) | 1313 (5.8%) | 52 (1.4%) |
| Health Care and Social Assistance | 1562 (7.2%) | 248 (5.6%) | 1629 (7.2%) | 181 (4.8%) |
| Information Services | 288 (1.3%) | 55 (1.2%) | 293 (1.3%) | 50 (1.3%) |
| Manufacturing [Ref.] | 3691 (17.0%) | 854 (19.1%) | 3808 (16.9%) | 737 (19.7%) |
| Personal Services | 295 (1.4%) | 146 (3.3%) | 299 (1.3%) | 142 (3.8%) |
| Professional, Scientific, Technical and Educational Services | 1811 (8.3%) | 797 (17.9%) | 1928 (8.6%) | 680 (18.1%) |
| Public Administration | 933 (4.3%) | 533 (11.9%) | 1042 (4.6%) | 424 (11.3%) |
| Transportation, Communications, and other Public Utilities | 1526 (7.0%) | 734 (16.4%) | 1606 (7.1%) | 654 (17.4%) |
| Wholesale and Retail Trade | 3851 (17.7%) | 315 (7.1%) | 3904 (17.4%) | 262 (7.0%) |
| Missing | 111 (0.5%) | 10 (0.2%) | 115 (0.5%) | 6 (0.2%) |
| **Region of residence** |  |  |  |  |
| Midsouth Atlantic | 7287 (33.5%) | 1584 (35.5%) | 7587 (33.7%) | 1284 (34.2%) |
| Midwest | 5238 (24.1%) | 1241 (27.8%) | 5392 (24.0%) | 1087 (29.0%) |
| New England [Ref.] | 652 (3.0%) | 142 (3.2%) | 658 (2.9%) | 136 (3.6%) |
| Pacific | 2795 (12.8%) | 744 (16.7%) | 2870 (12.8%) | 669 (17.8%) |
| South | 4403 (20.2%) | 548 (12.3%) | 4540 (20.2%) | 411 (11.0%) |
| West | 1317 (6.0%) | 176 (3.9%) | 1348 (6.0%) | 145 (3.9%) |
| Other | 79 (0.4%) | 29 (0.6%) | 91 (0.4%) | 17 (0.5%) |
| **Recruitment Year** |  |  |  |  |
| 2001 - 2004 | 9659 (44.4%) | 2398 (53.7%) | 9969 (44.3%) | 2088 (55.7%) |
| 2005 - 2009 | 4257 (19.6%) | 776 (17.4%) | 4359 (19.4%) | 674 (18.0%) |
| 2010 - 2014 | 2550 (11.7%) | 404 (9.1%) | 2638 (11.7%) | 316 (8.4%) |
| 2015 - 2021 | 5305 (24.4%) | 886 (19.8%) | 5520 (24.5%) | 671 (17.9%) |

### Missing Data and weights

Missing covariate data for parametric g-formula were multiple imputed using a random forest approach. We generated 5 imputed datasets using 30 iterations to ensure convergence, imputations were pooled using Rubin’s rules ^38^ to produce pooled estimates and standard errors that account for both within- and between-imputation variability (Supplementary S5-S12). All analyses incorporated longitudinal individual sampling weights to ensure national representativeness.

### Analytic Strategy

The analysis proceeded in two phases: an exploratory causal discovery phase and a confirmatory modeling phase.

#### Exploratory Causal Discovery

A fundamental step in modeling an effect is understanding how covariates relate to the primary outcome under study. This step is often overlooked, and adjusting for all covariates without distinguishing colliders, confounders or mediators may distort the effect^39^. Given the limited prior work and many covariates, we adopted a hybrid approach combining expert judgment with structure-learning algorithms^40,41^. To reduce complexity (both conceptual and computational) for later parametric g-formula modeling, time-varying covariates were lagged by just one wave (lag1). For causal discovery only we selected three cross-sectional time points. For each cut we ran in parallel 1) a Peter-Clark (PC) skeleton search^42^ using the CG-LRT (conditional Gaussian likelihood-ratio) test and 2) an expert knowledge encoding step specifying forbidden/required relations (which includes our main hypothesis). Outputs of these steps were combined using the Fast Greedy Equivalence Search (FGES) algorithm^43^ with the m-separation score to produce a DAG (Directed Acyclic Graph) per cut. We retained edges present in at least two of the three DAGs, adjudicated remaining discrepancies by expert review, and from this consensus DAG identified minimally sufficient adjustment sets to estimate total and direct effects.

#### Confirmatory Statistical Modeling

We choose parametric g-formula because it explicitly models the full joint distribution of longitudinal confounders and treatments, efficiently recovering causal effects under time-varying confounding. For observational data, such as PSID study, this method is appropriate to estimate the causal effects of union membership or union presence at the company-level on mental or self-reported health. Because exposure is not randomly assigned in observational studies, associations between exposures and outcomes may be driven by confounders that jointly influence both (e.g., gender, state or industry). The g-formula fits outcome and covariates models and then simulates counterfactual outcomes under different exposure levels (all-exposed vs. all-unexposed). and marginalizes over the observed covariate distribution to provide a population-average treatment effect that accounts for measured confounding and thereby emulates a randomized controlled trial. Importantly, this approach relies on the assumptions of no unmeasured confounding, correct model specification, and positivity (each level of the exposure being possible for all strata of covariates).

Guided by the DAGs obtained during the exploratory phase, we fitted eight outcome models in total (Supplementary S1-S4). For each combination of exposure (union membership and union presence in the workplace) and outcome (mental health and self-reported health) we fitted separated models to estimate the total (adjust confounders and excluding mediators) and direct effect (adjusting for confounders and mediators)

We estimated mean ratios for the continuous outcomes using parametric g-formula. We performed 1000 bootstrap iterations^44^, each simulating a sample size equal to the original dataset, to derive standard errors and 95 % confidence intervals. Analyses were run on the imputed datasets, and results were pooled using Rubin’s rules ^38^.

### Software

Analyses were performed in R (version 4.5.0)^45^ and Python (version 3.12.7)^46^. The panel dataset was constructed using the psidR package (v2.3)^47^. Data cleaning and visualization used tidyverse (v2.0)^48^. Causal discovery was perfomed with TETRAD GUI (v7.6.8)^49^, and DAGs adjusted with DAGitty (v3.1)^50^. Multiple imputation used the miceforest Python library (v5.7.0)^51^ with a random forest engine. The parametric g-formula was implemented using pygformula (v1.1.6)^52^.

## Results

### Descriptive statistics

Figure 1 shows time trends (2001 – 2021) in union membership and union presence (top panels) and mental health and self-reported health (bottom panels). Both union membership and presence have declined over the selected period, with a steadier decline in membership (17% in 2001 versus 11% in 2021) than for presence (19% versus 14%). Mental health remained relatively stable between 2001 and 2019 albeit with some variations between 2008-2015 and 2015-2019 and a sharp increase (i.e., a decline in mental health) in 2021 during the COVID-19 pandemic. By contrast, self-reported health means have constantly increased over this 20-year window starting around a value of 2.2 in 2001 and ending with a value of 2.45 in 2021 indicating a steady decline in self-reported health.

**Figure 1.**
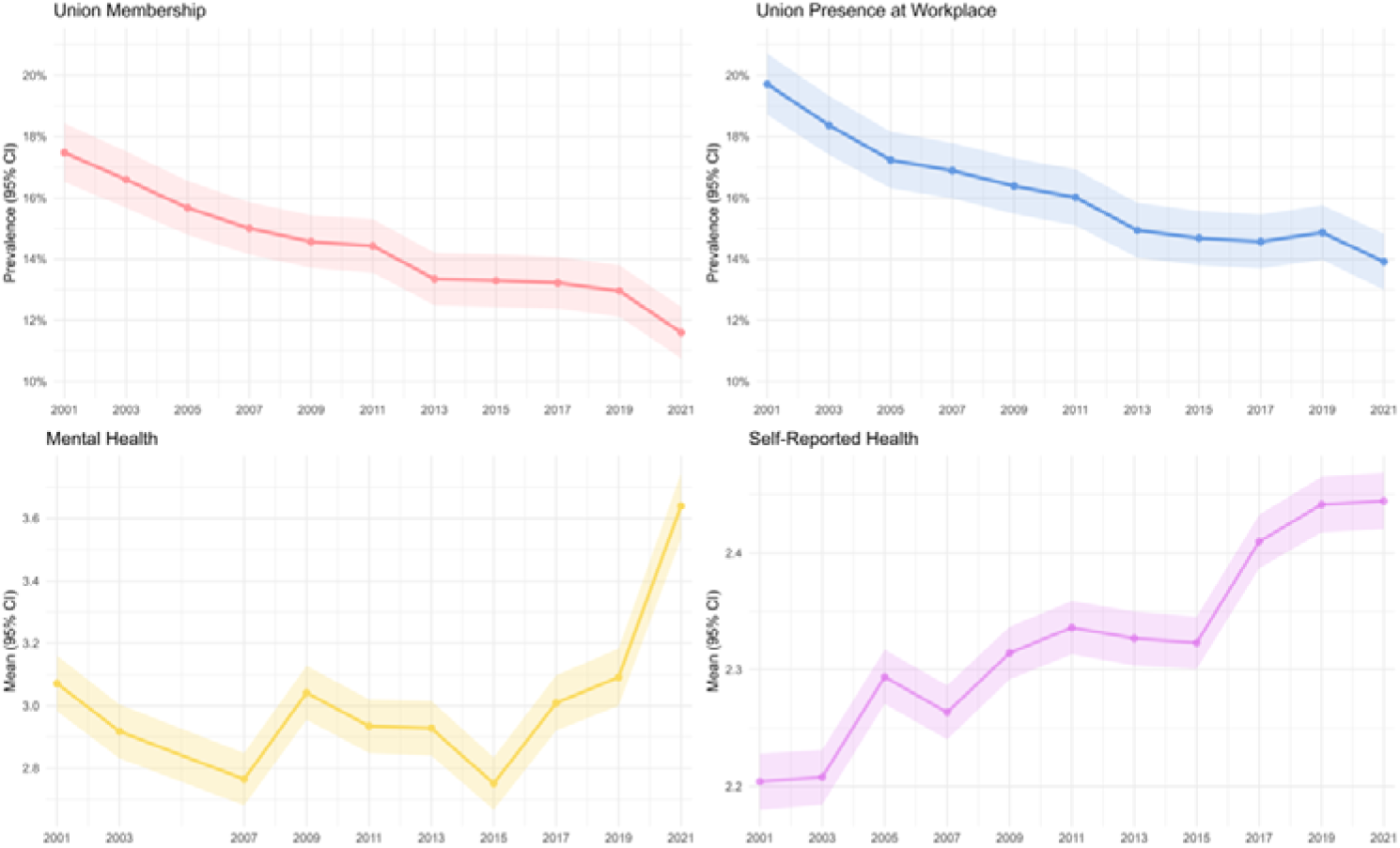
Descriptive statistics on mental health, self-reported health, union membership and workplace union presence (2001-2021)

**Table 1** reports baseline characteristics by union presence and union membership (N=26,235). Overall, 17% reported union presence at their workplace and 14% were union members. Compared with participants in non-unionized workplaces, those in unionized workplaces were slightly older and more likely to be men. Age distribution differed modestly: mid-career adults aged 35–54 years were more common among those with union presence, whereas younger workers (aged 18–34 years) predominated in non-union workplaces. The racial composition also varied, with a smaller proportion of white participants in unionized settings (49%) compared with non-union settings (56%).

Socioeconomic indicators suggested that workers in unionized workplaces had higher mean equivalized family income (US$ 42,300 vs 41,200) and a greater prevalence of home ownership (61% vs 52%). Household size was marginally larger among those with union presence. Educational attainment showed few differences overall, although non-degree tertiary education was slightly more common among unionized participants.

Industry and regional distributions showed the clearest contrasts. Union presence and membership were concentrated in manufacturing, transportation, public administration, and professional or technical services, and were relatively uncommon in accommodation, food service, and retail industries. Regionally, union presence was more frequent in the Pacific and Midwest and least common in the South. Participants from earlier recruitment waves (2001–04) were more likely to report union presence.

Health measures were broadly similar across groups. Mean self-reported general health scores were nearly identical, and mental health scores were slightly lower (indicating better mental wellbeing) among those with union presence.

### Causal associations of trade unions and mental health

Figure 2 displays directed acyclic graphs (DAGs) together with parametric g-formula results for mental health under total and direct adjustments. The upper panels use union membership as exposure and the lower use workplace union presence. Full DAGs and outcome model estimates are reported in Supplementary materials (Figures S1-S2; Tables S5-S8).

**Figure 2.**
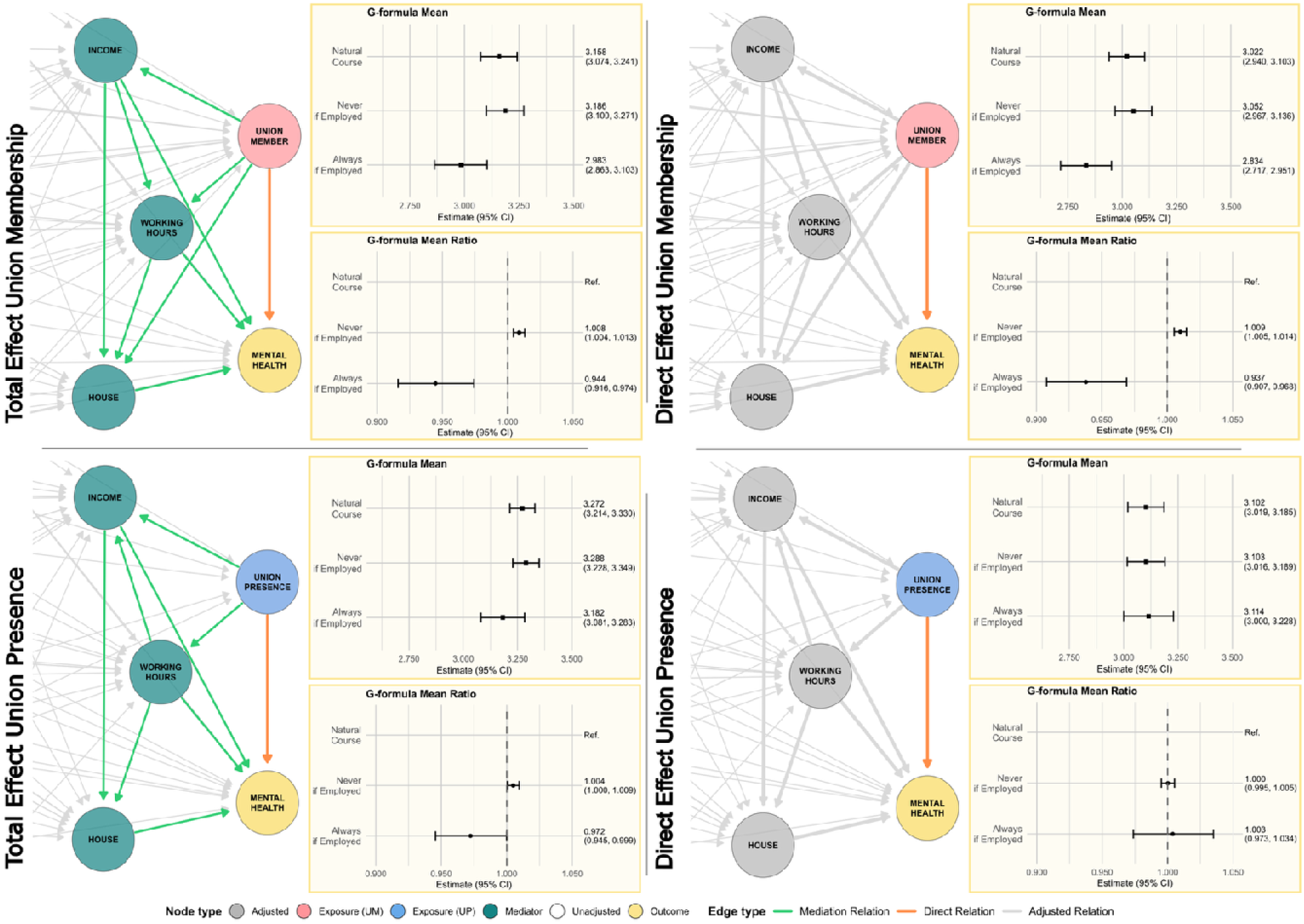
Total and direct effects of union membership and presence on mental health (Kesseler-6)

Under total adjustment (including mediators), with union membership as exposure, the parametric g-formula indicates that the counterfactual “never union member if employed” (labelled as Never if Employed) has a mean mental health score of 3.186 (95% CI 3.100–3.271) and a Mean Ratio (MR) of 1.008 (95% CI 1.004–1.103) relative to the natural course mean of 3.158 (95% CI 3.074–3.241), while the counterfactual “always union member if employed” (labelled as Always if Employed) yields a lower mean of 2.983 (95% CI 2.863–3.103; MR 0.944, 95% CI 0.916–0.974). When mediators are excluded (direct adjustment), “never union member if employed” (labelled Never if Employed) had a mean of 3.052 (95% CI 2.967–3.136; MR 1.009, 95% CI 1.005–1.104) versus the natural course mean of 3.022 (95% CI 2.940–3.103), and “always union member if employed” (labelled Always if Employed) a mean of 2.834 (95% CI 2.717–2.951; MR of 0.937, 95% CI 0.907–0.968).

For union presence in the workplace as the exposure, total adjustment “never presence in workplace if employed” (labelled as Never if Employed) produced a mean of 3.288 (95% CI 3.228–3.349) and a mean ratio (MR) of 1.004 (95% CI 1.000–1.009) compared with the natural course mean of 3.272 (95% CI 3.214–3.330), and “always presence in workplace if employed” (labelled Always if Employed) counterfactual in the same model has a mean of 3.182 (95% CI 3.081–3.283; MR 0.972, 95% CI 0.945–0.999). Under direct adjustment “never presence in workplace if employed” (labelled Never if Employed) returns a mean of 3.103 (95% CI 3.016–3.189; MR 1.000, 95% CI 0.995–1.005) versus the natural course mean of 3.102 (95% CI 3.019–3.185), and “always presence in workplace if employed” (label Always if Employed) mean is 3.114 (95% CI 3.000–3.228; MR 1.003, 95% CI 0.973–1.034) showing non-significant associations.

### Causal associations of trade unions and self-reported health

Figure 3 shows DAGs together with parametric g-formula results for the outcome self-reported health under total and direct adjustment sets. As in Figure 2, the upper panels use union membership as the exposure, and the lower panels use union presence in the workplace. Complete DAGs and outcome main model estimates are reported in Supplementary materials (Figures S3-S4; Tables S9-S12).

**Figure 3.**
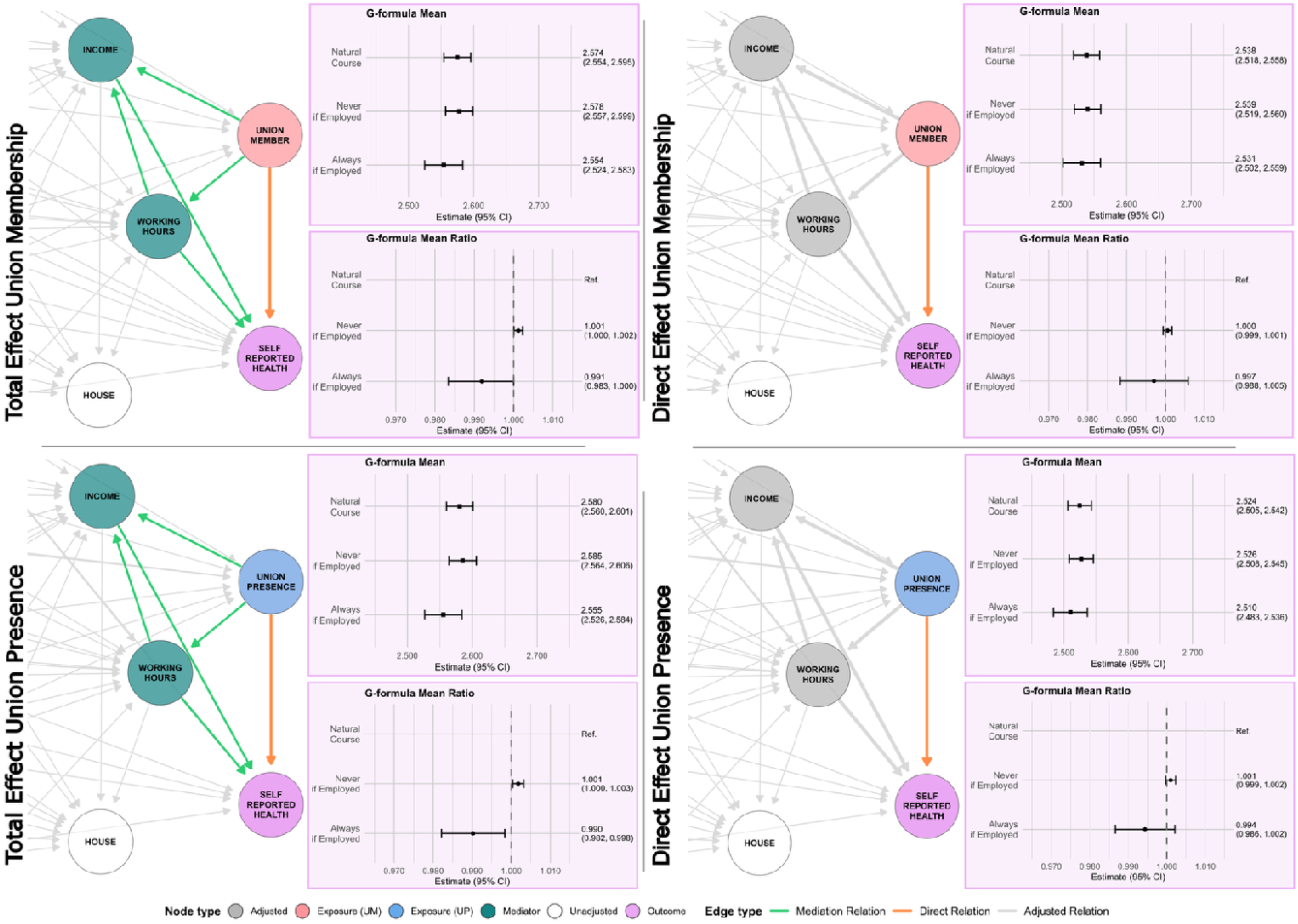
Total and direct effects of union membership and presence on Self-Rated Health.

When Union Membership is the exposure, and including mediators (total adjustment) the parametric g-formula estimated that the counterfactual “never union member if employed” (labelled as Never if Employed in Figure 3) had a mean self-reported health of 2.578 (95% CI 2.557–2.599) and a mean ratio (MR) of 1.001 (95% CI 1.000–1.002) relative to the natural course mean of 2.574 (95% CI 2.554–2.595), while the complementary scenario “always union member if employed” (labelled as Always if Employed) had a mean of 2.524 (95% CI 2.524–2.583; MR 0.991, 95% CI 0.983–1.000). Under direct adjustment “never union member if employed” (labelled Never if Employed) had a mean of 2.539 (95% CI 2.519–2.560; MR 1.000, 95% CI 0.999–1.001) versus natural course mean of 2.538 (95% CI 2.518–2.558), and always union member if employed” (labelled Always if Employed), had mean of 2.531 (95% CI 2.502–2.559), and a non-significant MR of 0.997 (95% CI 0.988–1.005),

For Union Presence in the Workplace, total adjustment model “never presence in workplace if employed” (labelled Never if Employed) had a mean of 2.585 (95% CI 2.564–2.606; MR 1.004, 95% CI 1.000–1.003) compared with the natural course mean of 2.580 (95% CI 2.560–2.601), and “always union presence in workplace if employed” (labelled as Always if Employed) mean of 2.555 (95% CI 2.526–2.584; MR 0.990, 95% CI 0.982–0.998). Under the direct adjustment the association was not statistically significant “never presence in workplace if employed” (Never if Employed) yields a mean of 2.526 (95% CI 2.508–2.545) and a mean ratio (MR) of 1.001 (95% CI 0.999–1.002) versus the natural course mean of 2.524 (95% CI 2.505–2.542), and “always presence in workplace if employed” (labelled Always if Employed) had mean 2.510 (95% CI 2.483–2.536; MR 0.994, 95% CI 0.986–1.002).

## Discussion

Using over two decades of US longitudinal data and applying the parametric g-formula to account for time-varying confounding, this study provides new causal evidence that trade unions influence workers’ health. It examines both union membership and workplace union presence, dimensions not previously addressed simultaneously.

We found that sustained union membership was associated with approximately 5% lower psychological distress (Kessler-6 scores), a meaningful population-level difference. This effect persisted after adjusting for income, working hours, and housing, indicating a direct effect. Constant workplace union presence also showed an association, but it attenuated after adjusting for these mediators. Improvements in self-reported health were primarily mediated by income and working-hours pathways. These results indicate two mechanisms. First, unions improve financial conditions and stabilize working hours, established health determinants ^53,54^. Second, union membership influences wellbeing through non-financial mechanisms like safer workplaces, collective voice, and reduced psychosocial strain. The persistence of associations after adjusting for income confirms that both mechanisms contribute.

Mental health responds quickly to psychosocial factors such as social belonging, perceived protection and collective voice ^55^, which membership provides. By contrast, self-reported health captures longer-term physical conditions ^57^, biological processes ^58^ and psychological conditions ^59^ that can improve more indirectly via socioeconomic mediators. Workplace union presence can extend these effects to non-members by shifting workplace norms and conditions. It is therefore not surprising that stronger, more consistent associations were found with mental health than with self-reported health.

Several limitations should be acknowledged. Despite the longitudinal design and causal modeling, residual confounding by unmeasured factors – such as workplace culture, local labor policies, or individual behaviors – cannot be ruled out; this was mitigated using DAG-based causal discovery and lagged covariates. Both exposures and outcomes were self-reported, introducing potential measurement error, which is unlikely to explain the consistent associations observed. Missing data were addressed via multiple imputation and pooled analysis, and models distinguished total from direct effects to avoid over-adjustment. Although our models identified mediators, we did not perform a formal causal mediation analysis. This limitation does not invalidate our results but means we cannot quantify each pathway’s contribution. Positivity violations were minimized by restricting analyses to appropriate covariate strata, and adjustments for industry, region, and employment status reduced bias from compositional differences.

The current landscape of labor rights therefore represents a significant public health concern. The systematic erosion of collective bargaining rights and freedom to unionize constitutes more than a labor market issue – it represents the dismantling of protective factors for worker health and wellbeing. Policies that strengthen workers’ rights to organize and bargain collectively function as evidence-based public health interventions. These include protecting union elections from employer interference, expanding bargaining rights to excluded workers, and ensuring meaningful enforcement of labor standards. By recognizing that attacks on unionization represent attacks on public health, we can better align health equity goals with policies that protect and expand workers’ collective power.

## Data Availability

The Panel Study of Income Dynamics (PSID) are freely available at: https://simba.isr.umich.edu/data/data.aspx

## Declarations

### Authors contribution (CRediT)

Conceptualization: JGH, JW, NH

Data Curation: JGH

Formal Analysis: JGH

Funding Acquisition: JW

Investigation: JGH, JW

Methodology: JGH, JW, TK

Project Administration: JGH, JW, NH

Resources: JGH, JW

Software: JGH

Supervision: JW

Validation: JW

Visualization: JGH

Writing – Original Draft Preparation: JGH, JW

Writing – Review & Editing: JW, NH, TK

