## Supplementary files for "The causal effect of trade unions on workers’ health: A parametric g-formula approach using longitudinal data from the Panel Study of Income Dynamics (PSID)"

**Supplementary file S.1. DAG of the causal total and indirect effects of trade union membership on mental health**

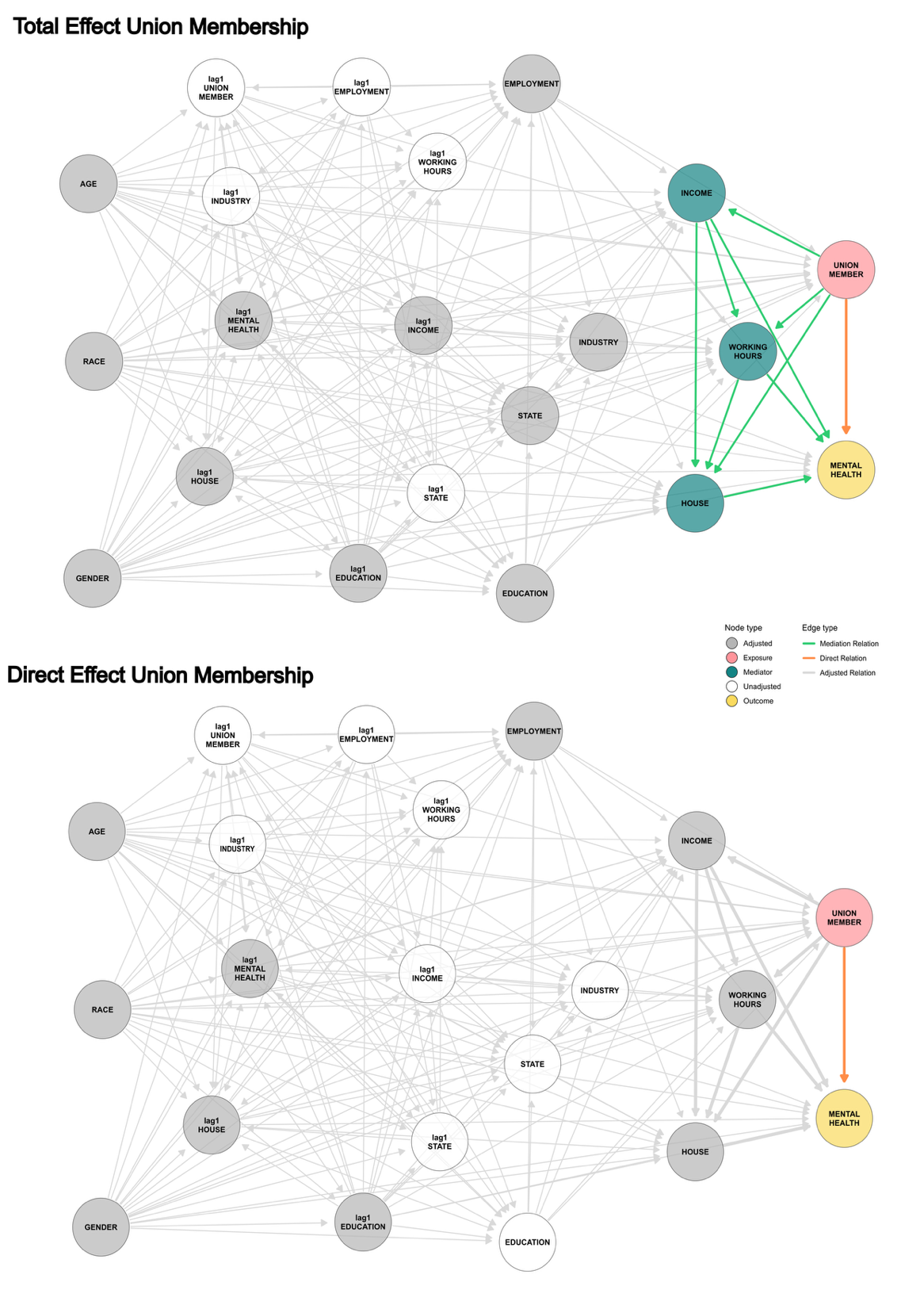

**Supplementary file S.2. DAG of the causal total and indirect effects of trade union presence and mental health**

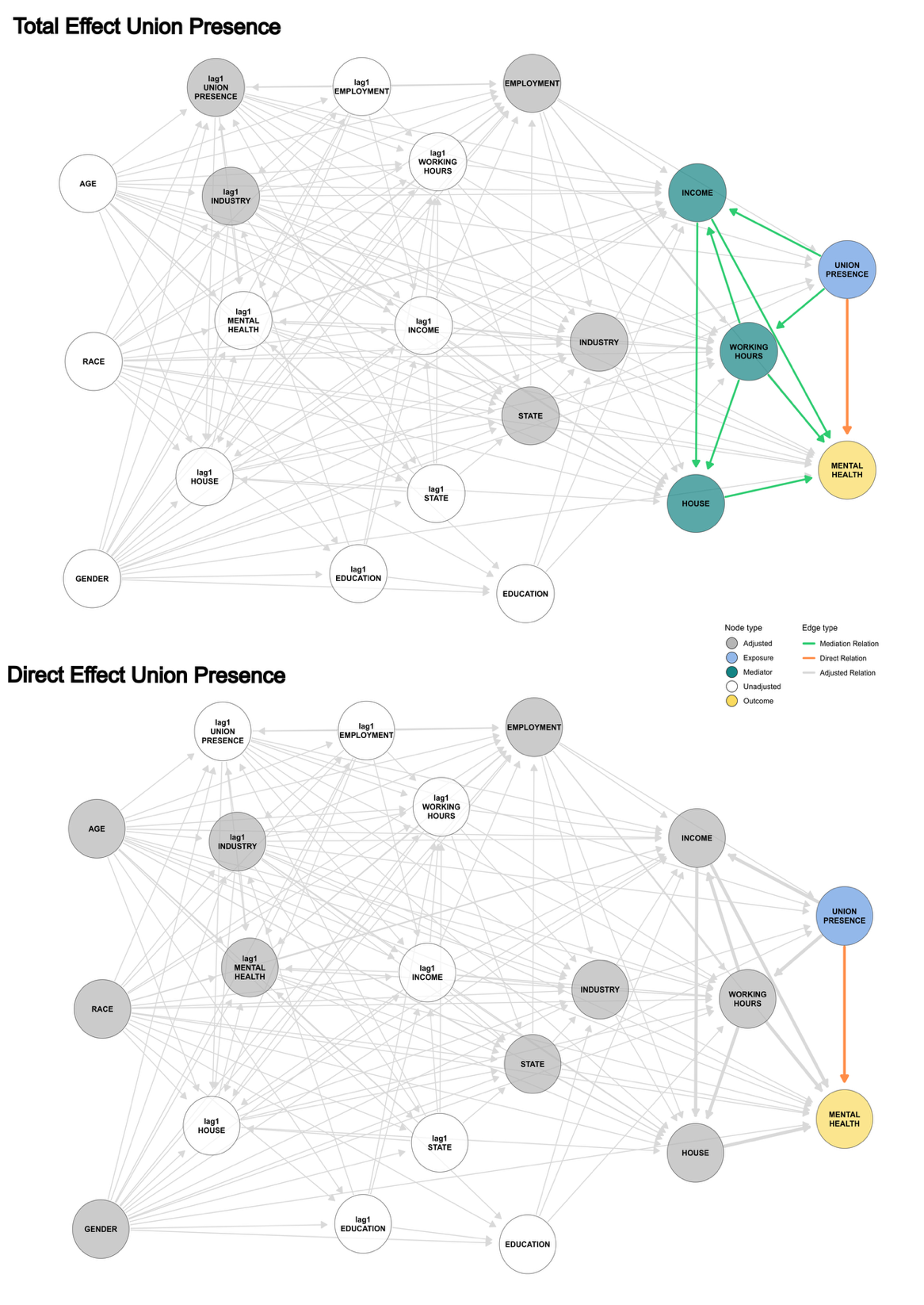

**Supplementary file S.3. DAG of the causal total and indirect effects of trade union membership on self-reported health**

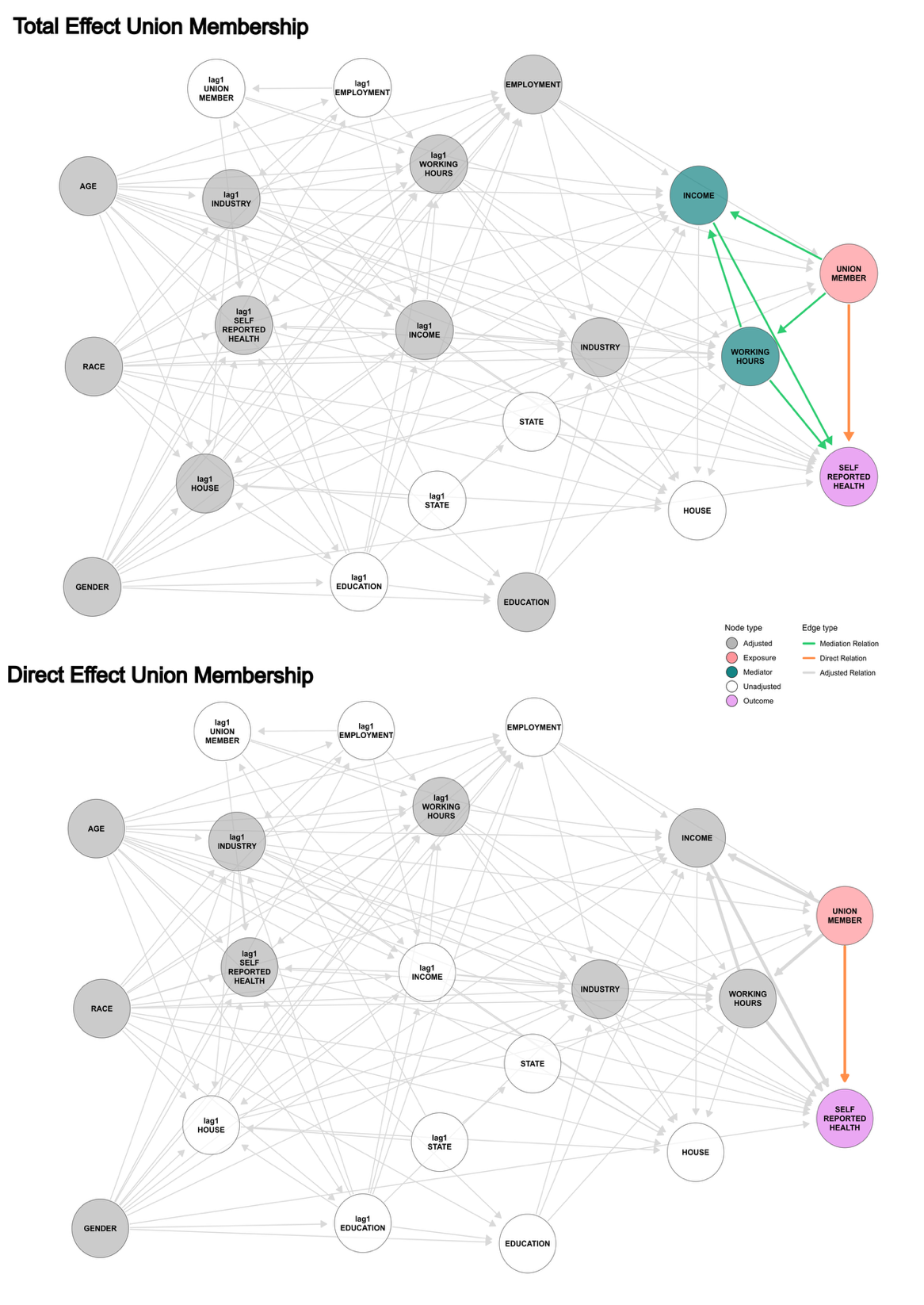

**Supplementary file S.4. DAG of the causal direct and indirect effects of trade union presence on self-reported health**

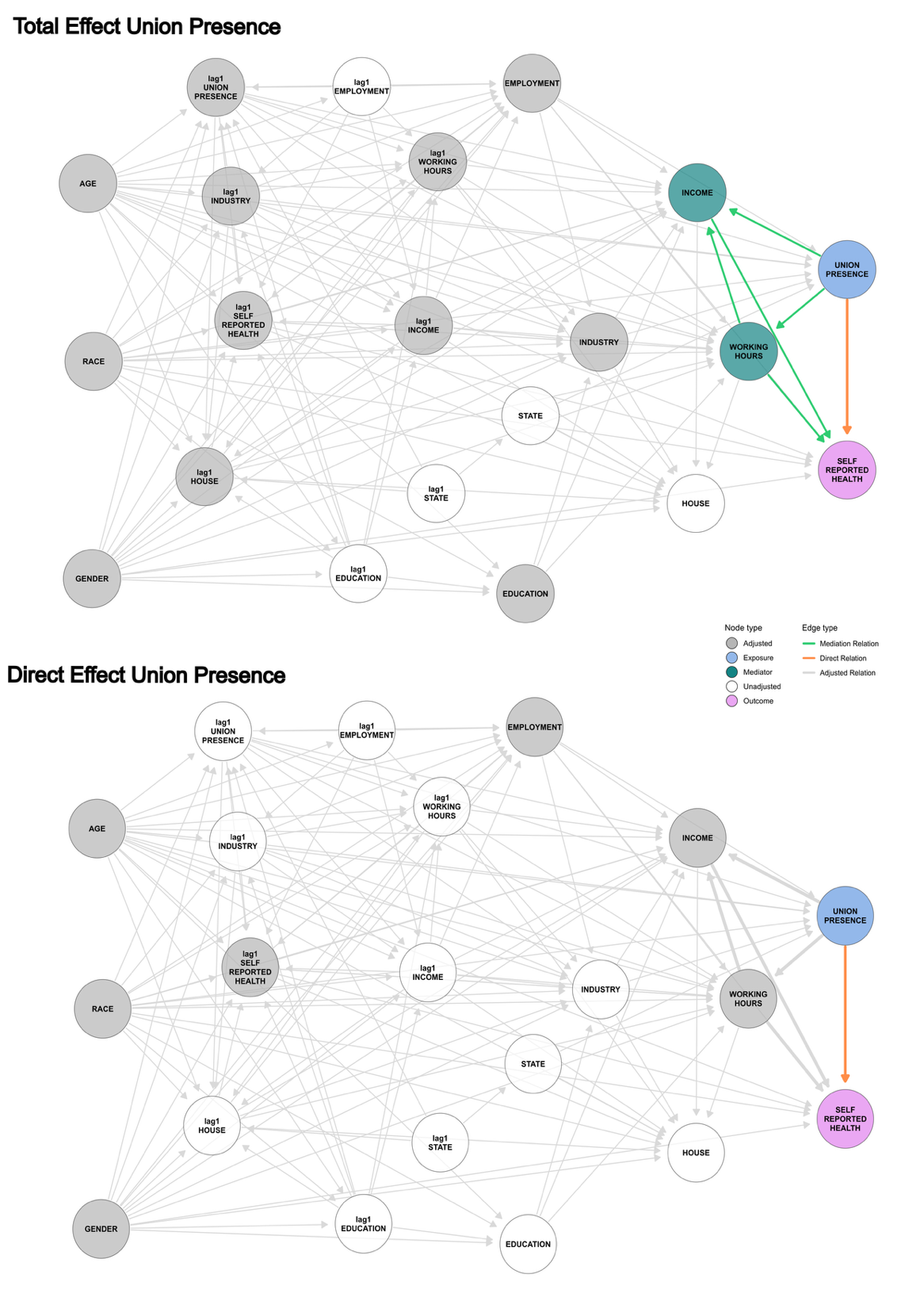

**Supplementary file S.5. Estimates and multiple imputation pooling statistics of the parametric g-formula outcome model of total effect of trade union membership on mental health**

|  | **Estimate** | **p-value** | **95% CI**  **lower** | **95% CI**  **lower** | **Pooled Std Err** | **Between-imputation var (B)** | **Within-imputation var (Ū)** | **Total var (T)** | **t-stat** | **n imputations (m)** |
| --- | --- | --- | --- | --- | --- | --- | --- | --- | --- | --- |
| Intercept | -45.451 | 0.000 | -52.614 | -38.288 | 3.655 | 0.013 | 13.341 | 13.357 | -12.436 | 5 |
| Union Membership | -0.075 | 0.015 | -0.136 | -0.014 | 0.031 | 0.000 | 0.001 | 0.001 | -2.426 | 5 |
| Age | -0.010 | 0.000 | -0.012 | -0.008 | 0.001 | 0.000 | 0.000 | 0.000 | -9.908 | 5 |
| Education University (Degree) | -0.213 | 0.000 | -0.300 | -0.125 | 0.045 | 0.000 | 0.002 | 0.002 | -4.753 | 5 |
| Education University (No Degree) | -0.161 | 0.000 | -0.240 | -0.081 | 0.041 | 0.000 | 0.002 | 0.002 | -3.973 | 5 |
| Education High School | 0.028 | 0.471 | -0.048 | 0.105 | 0.039 | 0.000 | 0.001 | 0.002 | 0.722 | 5 |
| Education Middle School | 0.217 | 0.016 | 0.041 | 0.393 | 0.090 | 0.000 | 0.007 | 0.008 | 2.419 | 5 |
| Education (NO U.S.) | 0.097 | 0.107 | -0.021 | 0.216 | 0.060 | 0.000 | 0.003 | 0.004 | 1.612 | 5 |
| Ind. Accommodation & Food serv. | 0.572 | 0.000 | 0.472 | 0.672 | 0.051 | 0.000 | 0.003 | 0.003 | 11.213 | 5 |
| Ind. Agriculture | -0.081 | 0.240 | -0.217 | 0.054 | 0.069 | 0.000 | 0.005 | 0.005 | -1.174 | 5 |
| Ind. Business & Repair serv. | 0.271 | 0.000 | 0.189 | 0.352 | 0.042 | 0.000 | 0.002 | 0.002 | 6.523 | 5 |
| Ind. Construction | -0.043 | 0.331 | -0.131 | 0.044 | 0.045 | 0.000 | 0.002 | 0.002 | -0.972 | 5 |
| Ind. Entertainment | 0.305 | 0.001 | 0.124 | 0.486 | 0.092 | 0.000 | 0.008 | 0.009 | 3.306 | 5 |
| Ind. Finance | -0.024 | 0.639 | -0.124 | 0.076 | 0.051 | 0.000 | 0.002 | 0.003 | -0.469 | 5 |
| Ind. Health & Social serv. | 0.022 | 0.602 | -0.062 | 0.106 | 0.043 | 0.000 | 0.002 | 0.002 | 0.522 | 5 |
| Ind. Education & Scientific | -0.064 | 0.162 | -0.153 | 0.026 | 0.046 | 0.000 | 0.002 | 0.002 | -1.400 | 5 |
| Ind. Public Administration | -0.283 | 0.000 | -0.375 | -0.190 | 0.047 | 0.000 | 0.002 | 0.002 | -6.003 | 5 |
| Ind. Utilities | 0.037 | 0.386 | -0.047 | 0.122 | 0.043 | 0.000 | 0.002 | 0.002 | 0.867 | 5 |
| Ind. Retail | 0.131 | 0.000 | 0.058 | 0.203 | 0.037 | 0.000 | 0.001 | 0.001 | 3.547 | 5 |
| Race (White) | 0.219 | 0.000 | 0.174 | 0.264 | 0.023 | 0.000 | 0.001 | 0.001 | 9.467 | 5 |
| Gender | 0.337 | 0.000 | 0.286 | 0.388 | 0.026 | 0.000 | 0.001 | 0.001 | 12.936 | 5 |
| Region South | 0.297 | 0.000 | 0.179 | 0.414 | 0.060 | 0.000 | 0.004 | 0.004 | 4.942 | 5 |
| Region Midwest | 0.225 | 0.000 | 0.111 | 0.339 | 0.058 | 0.000 | 0.003 | 0.003 | 3.877 | 5 |
| Region Midsouth Atlantic | 0.106 | 0.063 | -0.006 | 0.218 | 0.057 | 0.000 | 0.003 | 0.003 | 1.856 | 5 |
| Region Pacific | 0.162 | 0.008 | 0.043 | 0.282 | 0.061 | 0.000 | 0.004 | 0.004 | 2.657 | 5 |
| Region West | 0.145 | 0.035 | 0.010 | 0.280 | 0.069 | 0.000 | 0.005 | 0.005 | 2.104 | 5 |
| Lag1 Education University (Degree) | 0.014 | 0.781 | -0.086 | 0.115 | 0.051 | 0.000 | 0.002 | 0.003 | 0.278 | 5 |
| Lag1 Education University (No Degree) | 0.009 | 0.864 | -0.090 | 0.108 | 0.050 | 0.000 | 0.002 | 0.003 | 0.171 | 5 |
| Lag1 Education High School | -0.054 | 0.262 | -0.149 | 0.041 | 0.048 | 0.000 | 0.002 | 0.002 | -1.127 | 5 |
| Lag1 Education Middle School | -0.279 | 0.013 | -0.498 | -0.060 | 0.111 | 0.002 | 0.010 | 0.012 | -2.519 | 5 |
| Lag1 Education (NO U.S.) | 0.022 | 0.818 | -0.174 | 0.218 | 0.095 | 0.003 | 0.005 | 0.009 | 0.233 | 5 |
| lag1 House Owns | -0.230 | 0.000 | -0.283 | -0.177 | 0.027 | 0.000 | 0.001 | 0.001 | -8.504 | 5 |
| Lag1 Equivalized Income | -2.233 | 0.000 | -2.382 | -2.084 | 0.076 | 0.001 | 0.005 | 0.006 | -29.567 | 5 |
| Lag1 Mental Health | 0.468 | 0.000 | 0.462 | 0.474 | 0.003 | 0.000 | 0.000 | 0.000 | 155.912 | 5 |
| Year | 0.024 | 0.000 | 0.020 | 0.028 | 0.002 | 0.000 | 0.000 | 0.000 | 12.141 | 5 |

**Supplementary file S.6. Estimates and multiple imputation pooling statistics of the parametric g-formula outcome model of direct effect of trade union membership on mental health**

|  | **Estimate** | **p-value** | **95% CI**  **lower** | **95% CI**  **lower** | **Pooled Std Err** | **Between-imputation var (B)** | **Within-imputation var (Ū)** | **Total var (T)** | **t-stat** | **n imputations (m)** |
| --- | --- | --- | --- | --- | --- | --- | --- | --- | --- | --- |
| Intercept | -16.300 | 0.000 | -23.370 | -9.231 | 3.607 | 0.202 | 12.765 | 13.008 | -4.520 | 5 |
| Union Membership | -0.071 | 0.019 | -0.130 | -0.012 | 0.030 | 0.000 | 0.001 | 0.001 | -2.354 | 5 |
| Age | -0.010 | 0.000 | -0.012 | -0.008 | 0.001 | 0.000 | 0.000 | 0.000 | -10.038 | 5 |
| House Owns | -0.450 | 0.000 | -0.505 | -0.395 | 0.028 | 0.000 | 0.001 | 0.001 | -15.961 | 5 |
| Equivalized Income | -4.461 | 0.000 | -4.853 | -4.068 | 0.200 | 0.000 | 0.040 | 0.040 | -22.284 | 5 |
| Race (White) | 0.364 | 0.000 | 0.321 | 0.408 | 0.022 | 0.000 | 0.000 | 0.000 | 16.511 | 5 |
| Gender | 0.251 | 0.000 | 0.199 | 0.302 | 0.026 | 0.000 | 0.001 | 0.001 | 9.587 | 5 |
| Working Hours | -0.208 | 0.041 | -0.406 | -0.009 | 0.101 | 0.000 | 0.010 | 0.010 | -2.048 | 5 |
| Lag1 Education University (Degree) | -0.879 | 0.000 | -0.948 | -0.811 | 0.035 | 0.000 | 0.001 | 0.001 | -25.418 | 5 |
| Lag1 Education University (No Degree) | -0.934 | 0.000 | -1.000 | -0.867 | 0.033 | 0.000 | 0.001 | 0.001 | -27.900 | 5 |
| Lag1 Education High School | -0.893 | 0.000 | -0.957 | -0.829 | 0.032 | 0.000 | 0.001 | 0.001 | -27.680 | 5 |
| Lag1 Education Middle School | 0.214 | 0.062 | -0.012 | 0.440 | 0.108 | 0.004 | 0.006 | 0.012 | 1.981 | 5 |
| Lag1 Education (NO U.S.) | -0.093 | 0.427 | -0.342 | 0.156 | 0.112 | 0.007 | 0.005 | 0.013 | -0.827 | 5 |
| lag1 House Owns | -0.265 | 0.000 | -0.324 | -0.206 | 0.030 | 0.000 | 0.001 | 0.001 | -8.838 | 5 |
| Lag1 Mental Health | 0.433 | 0.000 | 0.427 | 0.439 | 0.003 | 0.000 | 0.000 | 0.000 | 139.923 | 5 |
| Year | 0.011 | 0.000 | 0.007 | 0.015 | 0.002 | 0.000 | 0.000 | 0.000 | 5.654 | 5 |

**Supplementary file S.7. Estimates and multiple imputation pooling statistics of the parametric g-formula outcome model of total effect of trade union presence on workplace on mental health**

|  | **Estimate** | **p-value** | **95% CI**  **lower** | **95% CI**  **lower** | **Pooled Std Err** | **Between-imputation var (B)** | **Within-imputation var (Ū)** | **Total var (T)** | **t-stat** | **n imputations (m)** |
| --- | --- | --- | --- | --- | --- | --- | --- | --- | --- | --- |
| Intercept | -32.206 | 0.000 | -39.719 | -24.694 | 3.833 | 0.056 | 14.631 | 14.692 | -8.402 | 5 |
| Union Presence | -0.043 | 0.278 | -0.122 | 0.035 | 0.040 | 0.000 | 0.002 | 0.002 | -1.085 | 5 |
| Ind. Accommodation & Food serv. | 1.003 | 0.000 | 0.879 | 1.127 | 0.063 | 0.000 | 0.004 | 0.004 | 15.859 | 5 |
| Ind. Agriculture | 0.031 | 0.745 | -0.154 | 0.216 | 0.094 | 0.000 | 0.009 | 0.009 | 0.325 | 5 |
| Ind. Business & Repair serv. | 0.510 | 0.000 | 0.411 | 0.609 | 0.050 | 0.000 | 0.002 | 0.003 | 10.129 | 5 |
| Ind. Construction | 0.126 | 0.026 | 0.015 | 0.237 | 0.057 | 0.000 | 0.003 | 0.003 | 2.224 | 5 |
| Ind. Entertainment | 0.624 | 0.000 | 0.402 | 0.847 | 0.114 | 0.000 | 0.013 | 0.013 | 5.500 | 5 |
| Ind. Finance | 0.135 | 0.042 | 0.005 | 0.266 | 0.067 | 0.000 | 0.004 | 0.004 | 2.032 | 5 |
| Ind. Health & Social serv. | 0.276 | 0.000 | 0.169 | 0.383 | 0.055 | 0.000 | 0.003 | 0.003 | 5.050 | 5 |
| Ind. Education & Scientific | 0.023 | 0.699 | -0.092 | 0.137 | 0.058 | 0.000 | 0.003 | 0.003 | 0.386 | 5 |
| Ind. Public Administration | -0.336 | 0.000 | -0.467 | -0.205 | 0.067 | 0.000 | 0.004 | 0.004 | -5.019 | 5 |
| Ind. Utilities | 0.074 | 0.181 | -0.034 | 0.182 | 0.055 | 0.000 | 0.003 | 0.003 | 1.339 | 5 |
| Ind. Retail | 0.313 | 0.000 | 0.227 | 0.398 | 0.044 | 0.000 | 0.002 | 0.002 | 7.186 | 5 |
| Lag1 Union Presence | 0.030 | 0.495 | -0.056 | 0.116 | 0.044 | 0.000 | 0.002 | 0.002 | 0.683 | 5 |
| Lag1 Ind. Accommodation & Food serv. | 0.103 | 0.137 | -0.033 | 0.240 | 0.069 | 0.000 | 0.005 | 0.005 | 1.489 | 5 |
| Lag1 Ind. Agriculture | -0.155 | 0.139 | -0.360 | 0.050 | 0.105 | 0.000 | 0.011 | 0.011 | -1.480 | 5 |
| Lag1 Ind. Business & Repair serv. | -0.203 | 0.000 | -0.307 | -0.098 | 0.053 | 0.000 | 0.003 | 0.003 | -3.808 | 5 |
| Lag1 Ind. Construction | -0.174 | 0.004 | -0.292 | -0.056 | 0.060 | 0.000 | 0.004 | 0.004 | -2.880 | 5 |
| Lag1 Ind. Entertainment | -0.332 | 0.013 | -0.592 | -0.071 | 0.133 | 0.001 | 0.017 | 0.018 | -2.494 | 5 |
| Lag1 Ind. Finance | -0.475 | 0.000 | -0.613 | -0.337 | 0.070 | 0.000 | 0.005 | 0.005 | -6.741 | 5 |
| Lag1 Ind. Health & Social serv. | -0.414 | 0.000 | -0.528 | -0.301 | 0.058 | 0.000 | 0.003 | 0.003 | -7.163 | 5 |
| Lag1 Ind. Education & Scientific | -0.525 | 0.000 | -0.645 | -0.405 | 0.061 | 0.000 | 0.004 | 0.004 | -8.563 | 5 |
| Lag1 Ind. Public Administration | -0.425 | 0.000 | -0.565 | -0.285 | 0.072 | 0.000 | 0.005 | 0.005 | -5.937 | 5 |
| Lag1 Ind. Utilities | -0.178 | 0.003 | -0.294 | -0.063 | 0.059 | 0.000 | 0.003 | 0.003 | -3.022 | 5 |
| Lag1 Ind. Retail | -0.224 | 0.000 | -0.309 | -0.139 | 0.044 | 0.000 | 0.002 | 0.002 | -5.147 | 5 |
| Region South | 0.488 | 0.000 | 0.361 | 0.615 | 0.065 | 0.000 | 0.004 | 0.004 | 7.506 | 5 |
| Region Midwest | 0.384 | 0.000 | 0.261 | 0.508 | 0.063 | 0.000 | 0.004 | 0.004 | 6.099 | 5 |
| Region Midsouth Atlantic | 0.141 | 0.023 | 0.019 | 0.262 | 0.062 | 0.000 | 0.004 | 0.004 | 2.272 | 5 |
| Region Pacific | 0.263 | 0.000 | 0.132 | 0.394 | 0.067 | 0.000 | 0.004 | 0.004 | 3.923 | 5 |
| Region West | 0.262 | 0.000 | 0.115 | 0.409 | 0.075 | 0.000 | 0.006 | 0.006 | 3.492 | 5 |
| Year | 0.017 | 0.000 | 0.013 | 0.021 | 0.002 | 0.000 | 0.000 | 0.000 | 8.656 | 5 |

**Supplementary file S.8. Estimates and multiple imputation pooling statistics of the parametric g-formula outcome model of direct effect of trade union presence on workplace on mental health**

|  | **Estimate** | **p-value** | **95% CI**  **lower** | **95% CI**  **lower** | **Pooled Std Err** | **Between-imputation var (B)** | **Within-imputation var (Ū)** | **Total var (T)** | **t-stat** | **n imputations (m)** |
| --- | --- | --- | --- | --- | --- | --- | --- | --- | --- | --- |
| Intercept | -5.419 | 0.130 | -12.435 | 1.597 | 3.580 | 0.097 | 12.696 | 12.813 | -1.514 | 5 |
| Union Presence | 0.030 | 0.308 | -0.027 | 0.086 | 0.029 | 0.000 | 0.001 | 0.001 | 1.019 | 5 |
| Gender | 0.283 | 0.000 | 0.232 | 0.334 | 0.026 | 0.000 | 0.001 | 0.001 | 10.886 | 5 |
| Age | -0.011 | 0.000 | -0.013 | -0.009 | 0.001 | 0.000 | 0.000 | 0.000 | -11.366 | 5 |
| Race (White) | 0.344 | 0.000 | 0.299 | 0.390 | 0.023 | 0.000 | 0.001 | 0.001 | 14.947 | 5 |
| Ind. Accommodation & Food serv. | 0.897 | 0.000 | 0.782 | 1.012 | 0.059 | 0.000 | 0.003 | 0.003 | 15.270 | 5 |
| Ind. Agriculture | 0.309 | 0.000 | 0.137 | 0.481 | 0.088 | 0.000 | 0.007 | 0.008 | 3.527 | 5 |
| Ind. Business & Repair serv. | 0.692 | 0.000 | 0.598 | 0.787 | 0.048 | 0.000 | 0.002 | 0.002 | 14.451 | 5 |
| Ind. Construction | 0.343 | 0.000 | 0.240 | 0.446 | 0.053 | 0.000 | 0.003 | 0.003 | 6.512 | 5 |
| Ind. Entertainment | 0.738 | 0.000 | 0.486 | 0.990 | 0.125 | 0.004 | 0.011 | 0.016 | 5.912 | 5 |
| Ind. Finance | 0.514 | 0.000 | 0.389 | 0.640 | 0.064 | 0.000 | 0.003 | 0.004 | 8.086 | 5 |
| Ind. Health & Social serv. | 0.496 | 0.000 | 0.397 | 0.595 | 0.050 | 0.000 | 0.002 | 0.003 | 9.855 | 5 |
| Ind. Education & Scientific | 0.537 | 0.000 | 0.429 | 0.644 | 0.055 | 0.000 | 0.003 | 0.003 | 9.774 | 5 |
| Ind. Public Administration | 0.147 | 0.018 | 0.025 | 0.269 | 0.062 | 0.000 | 0.004 | 0.004 | 2.370 | 5 |
| Ind. Utilities | 0.479 | 0.000 | 0.379 | 0.580 | 0.051 | 0.000 | 0.003 | 0.003 | 9.355 | 5 |
| Ind. Retail | 0.533 | 0.000 | 0.451 | 0.615 | 0.042 | 0.000 | 0.002 | 0.002 | 12.830 | 5 |
| Lag1 Union Presence | -1.082 | 0.000 | -1.209 | -0.956 | 0.065 | 0.000 | 0.004 | 0.004 | -16.777 | 5 |
| Lag1 Ind. Accommodation & Food serv. | -0.927 | 0.000 | -1.115 | -0.738 | 0.096 | 0.000 | 0.009 | 0.009 | -9.656 | 5 |
| Lag1 Ind. Agriculture | -0.965 | 0.000 | -1.065 | -0.865 | 0.051 | 0.000 | 0.002 | 0.003 | -18.926 | 5 |
| Lag1 Ind. Business & Repair serv. | -0.878 | 0.000 | -0.987 | -0.769 | 0.056 | 0.000 | 0.003 | 0.003 | -15.763 | 5 |
| Lag1 Ind. Construction | -1.123 | 0.000 | -1.407 | -0.840 | 0.141 | 0.005 | 0.014 | 0.020 | -7.959 | 5 |
| Lag1 Ind. Entertainment | -1.072 | 0.000 | -1.209 | -0.936 | 0.069 | 0.001 | 0.004 | 0.005 | -15.485 | 5 |
| Lag1 Ind. Finance | -1.118 | 0.000 | -1.223 | -1.013 | 0.054 | 0.000 | 0.003 | 0.003 | -20.822 | 5 |
| Lag1 Ind. Health & Social serv. | -1.227 | 0.000 | -1.338 | -1.117 | 0.056 | 0.000 | 0.003 | 0.003 | -21.796 | 5 |
| Lag1 Ind. Education & Scientific | -0.886 | 0.000 | -1.014 | -0.759 | 0.065 | 0.000 | 0.004 | 0.004 | -13.662 | 5 |
| Lag1 Ind. Public Administration | -0.867 | 0.000 | -0.973 | -0.761 | 0.054 | 0.000 | 0.003 | 0.003 | -16.054 | 5 |
| Lag1 Ind. Utilities | -1.011 | 0.000 | -1.094 | -0.927 | 0.043 | 0.000 | 0.002 | 0.002 | -23.752 | 5 |
| Region South | 0.301 | 0.000 | 0.183 | 0.418 | 0.060 | 0.000 | 0.004 | 0.004 | 5.009 | 5 |
| Region Midwest | 0.195 | 0.001 | 0.081 | 0.309 | 0.058 | 0.000 | 0.003 | 0.003 | 3.359 | 5 |
| Region Midsouth Atlantic | 0.113 | 0.052 | -0.001 | 0.226 | 0.058 | 0.000 | 0.003 | 0.003 | 1.941 | 5 |
| Region Pacific | 0.146 | 0.017 | 0.027 | 0.266 | 0.061 | 0.000 | 0.004 | 0.004 | 2.396 | 5 |
| Region West | 0.153 | 0.026 | 0.018 | 0.289 | 0.069 | 0.000 | 0.005 | 0.005 | 2.224 | 5 |
| Lag1 Mental Health | 0.424 | 0.000 | 0.418 | 0.430 | 0.003 | 0.000 | 0.000 | 0.000 | 140.607 | 5 |
| Working Hours | -0.153 | 0.135 | -0.353 | 0.048 | 0.102 | 0.000 | 0.010 | 0.010 | -1.493 | 5 |
| Equivalized Income | -4.440 | 0.000 | -4.832 | -4.048 | 0.200 | 0.000 | 0.040 | 0.040 | -22.186 | 5 |
| Housing Owns | -0.586 | 0.000 | -0.633 | -0.539 | 0.024 | 0.000 | 0.001 | 0.001 | -24.420 | 5 |
| Year | 0.006 | 0.005 | 0.002 | 0.010 | 0.002 | 0.000 | 0.000 | 0.000 | 2.818 | 5 |

**Supplementary file S.9. Estimates and multiple imputation pooling statistics of the parametric g-formula outcome model of total effect of trade union membership on self-reported health**

|  | **Estimate** | **p-value** | **95% CI**  **lower** | **95% CI**  **lower** | **Pooled Std Err** | **Between-imputation var (B)** | **Within-imputation var (Ū)** | **Total var (T)** | **t-stat** | **n imputations (m)** |
| --- | --- | --- | --- | --- | --- | --- | --- | --- | --- | --- |
| Intercept | -15.847 | 0.000 | -17.495 | -14.200 | 0.841 | 0.001 | 0.706 | 0.707 | -18.852 | 5 |
| Union Membership | -0.001 | 0.943 | -0.014 | 0.013 | 0.007 | 0.000 | 0.000 | 0.000 | -0.071 | 5 |
| Age | 0.005 | 0.000 | 0.005 | 0.006 | 0.000 | 0.000 | 0.000 | 0.000 | 27.000 | 5 |
| Education (NO U.S.) | 0.041 | 0.056 | -0.001 | 0.082 | 0.020 | 0.000 | 0.000 | 0.000 | 2.067 | 5 |
| Education Middle School | 0.151 | 0.000 | 0.117 | 0.184 | 0.017 | 0.000 | 0.000 | 0.000 | 8.943 | 5 |
| Education High School | 0.006 | 0.605 | -0.020 | 0.033 | 0.012 | 0.000 | 0.000 | 0.000 | 0.535 | 5 |
| Education University (No Degree) | -0.061 | 0.001 | -0.090 | -0.032 | 0.013 | 0.000 | 0.000 | 0.000 | -4.700 | 5 |
| Education University (Degree) | -0.152 | 0.000 | -0.180 | -0.124 | 0.013 | 0.000 | 0.000 | 0.000 | -12.055 | 5 |
| Ind. Accommodation & Food serv. | 0.074 | 0.000 | 0.047 | 0.102 | 0.014 | 0.000 | 0.000 | 0.000 | 5.273 | 5 |
| Ind. Agriculture | 0.088 | 0.000 | 0.048 | 0.129 | 0.021 | 0.000 | 0.000 | 0.000 | 4.264 | 5 |
| Ind. Business & Repair serv. | -0.013 | 0.235 | -0.035 | 0.009 | 0.011 | 0.000 | 0.000 | 0.000 | -1.187 | 5 |
| Ind. Construction | -0.011 | 0.362 | -0.035 | 0.013 | 0.012 | 0.000 | 0.000 | 0.000 | -0.911 | 5 |
| Ind. Entertainment | 0.003 | 0.922 | -0.048 | 0.053 | 0.026 | 0.000 | 0.001 | 0.001 | 0.098 | 5 |
| Ind. Finance | -0.006 | 0.692 | -0.033 | 0.022 | 0.014 | 0.000 | 0.000 | 0.000 | -0.396 | 5 |
| Ind. Health & Social serv. | 0.020 | 0.094 | -0.003 | 0.044 | 0.012 | 0.000 | 0.000 | 0.000 | 1.675 | 5 |
| Ind. Education & Scientific | -0.012 | 0.383 | -0.038 | 0.015 | 0.013 | 0.000 | 0.000 | 0.000 | -0.872 | 5 |
| Ind. Public Administration | -0.029 | 0.047 | -0.057 | -0.000 | 0.014 | 0.000 | 0.000 | 0.000 | -1.992 | 5 |
| Ind. Utilities | 0.028 | 0.022 | 0.004 | 0.053 | 0.012 | 0.000 | 0.000 | 0.000 | 2.297 | 5 |
| Ind. Retail | 0.035 | 0.001 | 0.015 | 0.055 | 0.010 | 0.000 | 0.000 | 0.000 | 3.433 | 5 |
| Race (White) | -0.071 | 0.000 | -0.081 | -0.061 | 0.005 | 0.000 | 0.000 | 0.000 | -14.135 | 5 |
| Gender | 0.051 | 0.000 | 0.039 | 0.062 | 0.006 | 0.000 | 0.000 | 0.000 | 8.413 | 5 |
| Lag1 House Owns | -0.003 | 0.627 | -0.015 | 0.009 | 0.006 | 0.000 | 0.000 | 0.000 | -0.486 | 5 |
| Lag1 Equivalized Income | -1.442 | 0.000 | -1.515 | -1.369 | 0.037 | 0.000 | 0.001 | 0.001 | -38.788 | 5 |
| Lag1 Ind. Accommodation & Food serv. | -0.107 | 0.000 | -0.139 | -0.075 | 0.016 | 0.000 | 0.000 | 0.000 | -6.593 | 5 |
| Lag1 Ind. Agriculture | -0.015 | 0.518 | -0.061 | 0.030 | 0.023 | 0.000 | 0.001 | 0.001 | -0.647 | 5 |
| Lag1 Ind. Business & Repair serv. | -0.018 | 0.170 | -0.044 | 0.008 | 0.013 | 0.000 | 0.000 | 0.000 | -1.372 | 5 |
| Lag1 Ind. Construction | 0.001 | 0.969 | -0.027 | 0.028 | 0.014 | 0.000 | 0.000 | 0.000 | 0.038 | 5 |
| Lag1 Ind. Entertainment | -0.063 | 0.033 | -0.120 | -0.005 | 0.029 | 0.000 | 0.001 | 0.001 | -2.141 | 5 |
| Lag1 Ind. Finance | -0.010 | 0.556 | -0.041 | 0.022 | 0.016 | 0.000 | 0.000 | 0.000 | -0.588 | 5 |
| Lag1 Ind. Health & Social serv. | -0.045 | 0.001 | -0.073 | -0.018 | 0.014 | 0.000 | 0.000 | 0.000 | -3.195 | 5 |
| Lag1 Ind. Education & Scientific | -0.034 | 0.017 | -0.063 | -0.006 | 0.014 | 0.000 | 0.000 | 0.000 | -2.381 | 5 |
| Lag1 Ind. Public Administration | 0.014 | 0.386 | -0.018 | 0.047 | 0.017 | 0.000 | 0.000 | 0.000 | 0.868 | 5 |
| Lag1 Ind. Utilities | -0.013 | 0.356 | -0.041 | 0.015 | 0.014 | 0.000 | 0.000 | 0.000 | -0.924 | 5 |
| Lag1 Ind. Retail | -0.048 | 0.000 | -0.071 | -0.026 | 0.011 | 0.000 | 0.000 | 0.000 | -4.233 | 5 |
| Lag1 Self-Reported Health | 0.483 | 0.000 | 0.477 | 0.489 | 0.003 | 0.000 | 0.000 | 0.000 | 160.944 | 5 |
| Lag 1 Working Hours | -0.092 | 0.002 | -0.149 | -0.035 | 0.029 | 0.000 | 0.001 | 0.001 | -3.159 | 5 |
| Year | 0.009 | 0.000 | 0.009 | 0.009 | 0.000 | 0.000 | 0.000 | 0.000 | 45.000 | 5 |

**Supplementary file S.10. Estimates and multiple imputation pooling statistics of the parametric g-formula outcome model of direct effect of trade union membership on self-reported health**

|  | **Estimate** | **p-value** | **95% CI**  **lower** | **95% CI**  **lower** | **Pooled Std Err** | **Between-imputation var (B)** | **Within-imputation var (Ū)** | **Total var (T)** | **t-stat** | **n imputations (m)** |
| --- | --- | --- | --- | --- | --- | --- | --- | --- | --- | --- |
| Intercept | -2.304 | 0.008 | -4.015 | -0.593 | 0.873 | 0.009 | 0.751 | 0.762 | -2.639 | 5 |
| Union Membership | 0.013 | 0.107 | -0.003 | 0.029 | 0.008 | 0.000 | 0.000 | 0.000 | 1.612 | 5 |
| Age | 0.004 | 0.000 | 0.004 | 0.004 | 0.000 | 0.000 | 0.000 | 0.000 | 20.000 | 5 |
| Equivalized Income | -1.451 | 0.000 | -1.546 | -1.357 | 0.048 | 0.000 | 0.002 | 0.002 | -30.140 | 5 |
| Ind. Accommodation & Food serv. | 0.288 | 0.000 | 0.257 | 0.319 | 0.016 | 0.000 | 0.000 | 0.000 | 18.314 | 5 |
| Ind. Agriculture | 0.339 | 0.000 | 0.296 | 0.382 | 0.022 | 0.000 | 0.000 | 0.000 | 15.343 | 5 |
| Ind. Business & Repair serv. | 0.197 | 0.000 | 0.168 | 0.225 | 0.014 | 0.000 | 0.000 | 0.000 | 14.107 | 5 |
| Ind. Construction | 0.237 | 0.000 | 0.206 | 0.268 | 0.015 | 0.000 | 0.000 | 0.000 | 15.501 | 5 |
| Ind. Entertainment | 0.200 | 0.000 | 0.110 | 0.289 | 0.041 | 0.001 | 0.001 | 0.002 | 4.908 | 5 |
| Ind. Finance | 0.224 | 0.000 | 0.185 | 0.263 | 0.019 | 0.000 | 0.000 | 0.000 | 11.830 | 5 |
| Ind. Health & Social serv. | 0.239 | 0.000 | 0.208 | 0.269 | 0.015 | 0.000 | 0.000 | 0.000 | 15.979 | 5 |
| Ind. Education & Scientific | 0.241 | 0.000 | 0.210 | 0.272 | 0.015 | 0.000 | 0.000 | 0.000 | 15.663 | 5 |
| Ind. Public Administration | 0.206 | 0.000 | 0.170 | 0.243 | 0.018 | 0.000 | 0.000 | 0.000 | 11.433 | 5 |
| Ind. Utilities | 0.285 | 0.000 | 0.247 | 0.323 | 0.018 | 0.000 | 0.000 | 0.000 | 15.947 | 5 |
| Ind. Retail | 0.251 | 0.000 | 0.229 | 0.272 | 0.011 | 0.000 | 0.000 | 0.000 | 22.750 | 5 |
| Race (White) | -0.121 | 0.000 | -0.131 | -0.111 | 0.005 | 0.000 | 0.000 | 0.000 | -23.775 | 5 |
| gender | 0.084 | 0.000 | 0.072 | 0.096 | 0.006 | 0.000 | 0.000 | 0.000 | 13.961 | 5 |
| Working Hours | -0.069 | 0.006 | -0.118 | -0.020 | 0.025 | 0.000 | 0.001 | 0.001 | -2.739 | 5 |
| Lag1 Ind. Accommodation & Food serv. | -0.505 | 0.000 | -0.538 | -0.471 | 0.017 | 0.000 | 0.000 | 0.000 | -29.891 | 5 |
| Lag1 Ind. Agriculture | -0.404 | 0.000 | -0.453 | -0.355 | 0.025 | 0.000 | 0.001 | 0.001 | -16.164 | 5 |
| Lag1 Ind. Business & Repair serv. | -0.467 | 0.000 | -0.497 | -0.438 | 0.015 | 0.000 | 0.000 | 0.000 | -31.927 | 5 |
| Lag1 Ind. Construction | -0.440 | 0.000 | -0.471 | -0.408 | 0.016 | 0.000 | 0.000 | 0.000 | -27.739 | 5 |
| Lag1 Ind. Entertainment | -0.499 | 0.000 | -0.611 | -0.386 | 0.050 | 0.001 | 0.001 | 0.003 | -9.957 | 5 |
| Lag1 Ind. Finance | -0.503 | 0.000 | -0.542 | -0.464 | 0.019 | 0.000 | 0.000 | 0.000 | -25.950 | 5 |
| Lag1 Ind. Health & Social serv. | -0.499 | 0.000 | -0.530 | -0.467 | 0.016 | 0.000 | 0.000 | 0.000 | -31.689 | 5 |
| Lag1 Ind. Education & Scientific | -0.572 | 0.000 | -0.603 | -0.541 | 0.016 | 0.000 | 0.000 | 0.000 | -36.716 | 5 |
| Lag1 Ind. Public Administration | -0.491 | 0.000 | -0.528 | -0.454 | 0.019 | 0.000 | 0.000 | 0.000 | -26.487 | 5 |
| Lag1 Ind. Utilities | -0.472 | 0.000 | -0.513 | -0.432 | 0.019 | 0.000 | 0.000 | 0.000 | -24.432 | 5 |
| Lag1 Ind. Retail | -0.471 | 0.000 | -0.492 | -0.451 | 0.010 | 0.000 | 0.000 | 0.000 | -45.255 | 5 |
| Lag1 Self-Reported Health | 0.317 | 0.000 | 0.311 | 0.323 | 0.003 | 0.000 | 0.000 | 0.000 | 101.996 | 5 |
| Year | 0.003 | 0.000 | 0.002 | 0.003 | 0.000 | 0.000 | 0.000 | 0.000 | 42.667 | 5 |

**Supplementary file S.11. Estimates and multiple imputation pooling statistics of the parametric g-formula outcome model of total effect of trade union presence on workplace on self-reported health**

|  | **Estimate** | **p-value** | **95% CI**  **lower** | **95% CI**  **lower** | **Pooled Std Err** | **Between-imputation var (B)** | **Within-imputation var (Ū)** | **Total var (T)** | **t-stat** | **n imputations (m)** |
| --- | --- | --- | --- | --- | --- | --- | --- | --- | --- | --- |
| Intercept | -15.821 | 0.000 | -17.470 | -14.171 | 0.841 | 0.002 | 0.706 | 0.708 | -18.801 | 5 |
| Union Presence | 0.007 | 0.433 | -0.011 | 0.025 | 0.009 | 0.000 | 0.000 | 0.000 | 0.784 | 5 |
| Age | 0.005 | 0.000 | 0.005 | 0.006 | 0.000 | 0.000 | 0.000 | 0.000 | 27.000 | 5 |
| Education (NO U.S.) | 0.032 | 0.112 | -0.008 | 0.073 | 0.019 | 0.000 | 0.000 | 0.000 | 1.678 | 5 |
| Education Middle School | 0.151 | 0.000 | 0.110 | 0.192 | 0.020 | 0.000 | 0.000 | 0.000 | 7.591 | 5 |
| Education High School | 0.004 | 0.652 | -0.013 | 0.021 | 0.008 | 0.000 | 0.000 | 0.000 | 0.454 | 5 |
| Education University (No Degree) | -0.062 | 0.000 | -0.082 | -0.043 | 0.010 | 0.000 | 0.000 | 0.000 | -6.487 | 5 |
| Education University (Degree) | -0.152 | 0.000 | -0.173 | -0.131 | 0.010 | 0.000 | 0.000 | 0.000 | -14.815 | 5 |
| Ind. Accommodation & Food serv. | 0.068 | 0.000 | 0.040 | 0.096 | 0.014 | 0.000 | 0.000 | 0.000 | 4.792 | 5 |
| Ind. Agriculture | 0.082 | 0.000 | 0.040 | 0.124 | 0.021 | 0.000 | 0.000 | 0.000 | 3.868 | 5 |
| Ind. Business & Repair serv. | -0.019 | 0.094 | -0.042 | 0.003 | 0.012 | 0.000 | 0.000 | 0.000 | -1.678 | 5 |
| Ind. Construction | -0.016 | 0.205 | -0.041 | 0.009 | 0.013 | 0.000 | 0.000 | 0.000 | -1.268 | 5 |
| Ind. Entertainment | -0.000 | 0.998 | -0.050 | 0.050 | 0.026 | 0.000 | 0.001 | 0.001 | -0.002 | 5 |
| Ind. Finance | -0.012 | 0.447 | -0.041 | 0.018 | 0.015 | 0.000 | 0.000 | 0.000 | -0.760 | 5 |
| Ind. Health & Social serv. | 0.016 | 0.217 | -0.009 | 0.040 | 0.013 | 0.000 | 0.000 | 0.000 | 1.235 | 5 |
| Ind. Information | -0.052 | 0.014 | -0.094 | -0.011 | 0.021 | 0.000 | 0.000 | 0.000 | -2.467 | 5 |
| Ind. Education & Scientific | -0.018 | 0.186 | -0.045 | 0.009 | 0.014 | 0.000 | 0.000 | 0.000 | -1.322 | 5 |
| Ind. Public Administration | -0.036 | 0.016 | -0.066 | -0.007 | 0.015 | 0.000 | 0.000 | 0.000 | -2.402 | 5 |
| Ind. Utilities | 0.021 | 0.094 | -0.004 | 0.045 | 0.012 | 0.000 | 0.000 | 0.000 | 1.673 | 5 |
| Ind. Retail | 0.030 | 0.005 | 0.009 | 0.050 | 0.011 | 0.000 | 0.000 | 0.000 | 2.831 | 5 |
| Race (White) | -0.072 | 0.000 | -0.082 | -0.062 | 0.005 | 0.000 | 0.000 | 0.000 | -14.328 | 5 |
| Gender | 0.051 | 0.000 | 0.039 | 0.063 | 0.006 | 0.000 | 0.000 | 0.000 | 8.512 | 5 |
| Lag1 House Owns | -0.003 | 0.646 | -0.015 | 0.009 | 0.006 | 0.000 | 0.000 | 0.000 | -0.460 | 5 |
| Lag1 Equivalized Income | -1.436 | 0.000 | -1.511 | -1.362 | 0.038 | 0.000 | 0.001 | 0.001 | -37.749 | 5 |
| Lag1 Ind. Accommodation & Food serv. | -0.107 | 0.000 | -0.139 | -0.075 | 0.016 | 0.000 | 0.000 | 0.000 | -6.625 | 5 |
| Lag1 Ind. Agriculture | -0.016 | 0.485 | -0.062 | 0.029 | 0.023 | 0.000 | 0.001 | 0.001 | -0.698 | 5 |
| Lag1 Ind. Business & Repair serv. | -0.019 | 0.165 | -0.045 | 0.008 | 0.014 | 0.000 | 0.000 | 0.000 | -1.389 | 5 |
| Lag1 Ind. Construction | -0.000 | 1.000 | -0.028 | 0.028 | 0.014 | 0.000 | 0.000 | 0.000 | -0.000 | 5 |
| Lag1 Ind. Entertainment | -0.066 | 0.024 | -0.124 | -0.009 | 0.029 | 0.000 | 0.001 | 0.001 | -2.261 | 5 |
| Lag1 Ind. Finance | -0.012 | 0.475 | -0.044 | 0.020 | 0.016 | 0.000 | 0.000 | 0.000 | -0.715 | 5 |
| Lag1 Ind. Health & Social serv. | -0.048 | 0.001 | -0.076 | -0.019 | 0.015 | 0.000 | 0.000 | 0.000 | -3.250 | 5 |
| Lag1 Ind Information | 0.006 | 0.782 | -0.039 | 0.052 | 0.023 | 0.000 | 0.001 | 0.001 | 0.277 | 5 |
| Lag1 Ind. Education & Scientific | -0.033 | 0.035 | -0.063 | -0.002 | 0.015 | 0.000 | 0.000 | 0.000 | -2.112 | 5 |
| Lag1 Ind. Public Administration | 0.017 | 0.281 | -0.014 | 0.049 | 0.016 | 0.000 | 0.000 | 0.000 | 1.079 | 5 |
| Lag1 Ind. Utilities | -0.009 | 0.553 | -0.037 | 0.020 | 0.014 | 0.000 | 0.000 | 0.000 | -0.594 | 5 |
| Lag1 Ind. Retail | -0.050 | 0.000 | -0.073 | -0.028 | 0.012 | 0.000 | 0.000 | 0.000 | -4.364 | 5 |
| Lag1 Self-Reported Health | 0.483 | 0.000 | 0.477 | 0.489 | 0.003 | 0.000 | 0.000 | 0.000 | 160.604 | 5 |
| Lag1Union Presence | -0.018 | 0.065 | -0.038 | 0.001 | 0.010 | 0.000 | 0.000 | 0.000 | -1.847 | 5 |
| Lag1 Working Hours | -0.093 | 0.001 | -0.150 | -0.036 | 0.029 | 0.000 | 0.001 | 0.001 | -3.190 | 5 |
| Year | 0.009 | 0.000 | 0.009 | 0.009 | 0.000 | 0.000 | 0.000 | 0.000 | 149.000 | 5 |

**Supplementary file S.12. Estimates and multiple imputation pooling statistics of the parametric g-formula outcome model of direct effect of trade union presence on workplace on self-reported health**

|  | **Estimate** | **p-value** | **95% CI**  **lower** | **95% CI**  **lower** | **Pooled Std Err** | **Between-imputation var (B)** | **Within-imputation var (Ū)** | **Total var (T)** | **t-stat** | **n imputations (m)** |
| --- | --- | --- | --- | --- | --- | --- | --- | --- | --- | --- |
| Intercept | 4.370 | 0.000 | 2.657 | 6.083 | 0.874 | 0.000 | 0.764 | 0.764 | 5.000 | 5 |
| Union Presence | -0.000 | 0.988 | -0.014 | 0.014 | 0.007 | 0.000 | 0.000 | 0.000 | -0.015 | 5 |
| Age | 0.003 | 0.000 | 0.003 | 0.004 | 0.000 | 0.000 | 0.000 | 0.000 | 17.000 | 5 |
| Equivalized Income | -1.749 | 0.000 | -1.841 | -1.656 | 0.047 | 0.000 | 0.002 | 0.002 | -37.186 | 5 |
| Race (White) | -0.133 | 0.000 | -0.145 | -0.121 | 0.006 | 0.000 | 0.000 | 0.000 | -22.104 | 5 |
| Gender | 0.065 | 0.000 | 0.053 | 0.077 | 0.006 | 0.000 | 0.000 | 0.000 | 10.811 | 5 |
| Working Hours | -0.074 | 0.004 | -0.125 | -0.024 | 0.026 | 0.000 | 0.001 | 0.001 | -2.899 | 5 |
| Lag1 Self-Reported Health | 0.228 | 0.000 | 0.224 | 0.232 | 0.002 | 0.000 | 0.000 | 0.000 | 113.906 | 5 |
| Year | -0.001 | 0.003 | -0.001 | -0.000 | 0.000 | 0.000 | 0.000 | 0.000 | -3.000 | 5 |
